# Identification of driver genes for severe forms of COVID-19 in a deeply phenotyped young patient cohort

**DOI:** 10.1101/2021.06.21.21257822

**Authors:** Raphael Carapito, Richard Li, Julie Helms, Christine Carapito, Sharvari Gujja, Véronique Rolli, Raony Guimaraes, Jose Malagon-Lopez, Perrine Spinnhirny, Razieh Mohseninia, Aurélie Hirschler, Leslie Muller, Paul Bastard, Adrian Gervais, Qian Zhang, François Danion, Yvon Ruch, Maleka Schenck-Dhif, Olivier Collange, Thiên-Nga Chamaraux-Tran, Anne Molitor, Angélique Pichot, Alice Bernard, Ouria Tahar, Sabrina Bibi-Triki, Haiguo Wu, Nicodème Paul, Sylvain Mayeur, Annabel Larnicol, Géraldine Laumond, Julia Frappier, Sylvie Schmidt, Antoine Hanauer, Cécile Macquin, Tristan Stemmelen, Michael Simons, Xavier Mariette, Olivier Hermine, Samira Fafi-Kremer, Bernard Goichot, Bernard Drenou, Khaldoun Kuteifan, Julien Pottecher, Paul-Michel Mertes, Shweta Kailasan, M. Javad Aman, Elisa Pin, Peter Nilsson, Anne Thomas, Alain Viari, Damien Sanlaville, Francis Schneider, Jean Sibilia, Pierre-Louis Tharaux, Jean-Laurent Casanova, Yves Hansmann, Daniel Lidar, Mirjana Radosavljevic, Jeffrey R. Gulcher, Ferhat Meziani, Christiane Moog, Thomas W. Chittenden, Seiamak Bahram

## Abstract

The etiopathogenesis of severe COVID-19 remains unknown. Indeed given major confounding factors (age and co-morbidities), true drivers of this condition have remained elusive. Here, we employ an unprecedented multi-omics analysis, combined with artificial intelligence, in a young patient cohort where major co-morbidities have been excluded at the onset. Here, we established a three-tier cohort of individuals younger than 50 years without major comorbidities. These included 47 “critical” (in the ICU under mechanical ventilation) and 25 “non-critical” (in a noncritical care ward) COVID-19 patients as well as 22 healthy individuals. The analyses included whole-genome sequencing, whole-blood RNA sequencing, plasma and blood mononuclear cells proteomics, cytokine profiling and high-throughput immunophenotyping. An ensemble of machine learning, deep learning, quantum annealing and structural causal modeling led to key findings. Critical patients were characterized by exacerbated inflammation, perturbed lymphoid/myeloid compartments, coagulation and viral cell biology. Within a unique gene signature that differentiated critical from noncritical patients, several driver genes promoted severe COVID-19 among which the upregulated metalloprotease *ADAM9* was key. This gene signature was replicated in an independent cohort of 81 critical and 73 recovered COVID-19 patients, as were ADAM9 transcripts, soluble form and proteolytic activity. *Ex vivo* ADAM9 inhibition affected SARS-CoV-2 uptake and replication in human lung epithelial cells. In conclusion, within a young, otherwise healthy, COVID-19 cohort, we provide the landscape of biological perturbations *in vivo* where a unique gene signature differentiated critical from non-critical patients. The key driver, *ADAM9*, interfered with SARS-CoV-2 biology. A repositioning strategy for anti-ADAM9 therapeutic is feasible.

**One sentence summary:** Etiopathogenesis of severe COVID19 in a young patient population devoid of comorbidities.

## INTRODUCTION

Unlike many viral infections and most respiratory virus infections, COVID-19 is characterized by an extraordinarily complex and diversified spectrum of clinical manifestations, which results in the use of “syndemic” within, or *in lieu* of, pandemic (Horton, 2020). Indeed, upon infection with SARS-CoV-2, age-, sex-, and phenotype-matched individuals can be classified within four distinct groups, i.e., (1) asymptomatic individuals, (2) patients displaying influenza-like illnesses, (3) patients affected by respiratory dysfunction who eventually need an external oxygen supply, and (4) patients suffering from acute respiratory distress syndrome (ARDS) who need invasive mechanical ventilation in an intensive care unit (ICU). Even though the last group represents only a small fraction of COVID-19 patients, this group encompasses the most severe form of the disease and has an average case-fatality rate of approximately 25% (Quah et al., 2020). Despite intense investigation, the fundamental question of why the course of the disease shows such a marked difference in an otherwise, apparently indistinguishable set of individuals, remains despite key findings in discrete subpopulations (Van Der Made et al., 2020; Shelton et al., 2021; The Severe Covid-19 GWAS, 2020; Zhang et al., 2020), largely unanswered i.e. the exact pathophysiological mechanism governing the disease severity within a demographically and clinically homogeneous group of patients remains, for the majority of such patients, mostly unclear. To better understand this issue, high-resolution molecular analyses should be applied to well-defined cohorts of patients and controls where a maximum of confounding factors have been eliminated. These include notably older age as well as a number of co-morbidities – e.g., cerebrovascular disease, types 1 and 2 diabetes, chronic kidney disease, chronic obstructive pulmonary disease, heart conditions, etc. (CDC, 2021) – present in COVID-19 patients. This is the case for the three-tier cohort studied in this work.

Several studies have used single, or a restricted number of omics technologies to uncover key molecular processes associated with disease severity, usually in unfiltered severe COVID-19 patients. Systemic inflammation with high levels of acute-phase proteins (C reactive protein; CRP, serum amyloid A; SAA, calprotectin) (Silvin et al., 2020) and inflammatory cytokines, particularly interleukin (IL)-6 and IL-1β (Chen et al., 2020a; Giamarellos-Bourboulis et al., 2020; Lucas et al., 2020) has been found to be a hallmark of disease severity. In contrast, following an initial burst shortly after infection, the type I interferon (IFN) response is impaired at the RNA (Hadjadj et al., 2020) and plasma (Trouillet-Assant et al., 2020) levels. Severity was also correlated with profound immune dysregulations, including modifications in the myeloid compartment with increases in neutrophils (Meizlish et al., 2021; Schulte-Schrepping et al., 2020), decreases in nonclassical monocytes (Silvin et al., 2020) and dysregulation of macrophages (Giamarellos-Bourboulis et al., 2020; Shen et al., 2020). The lymphoid compartment is also modified by both a B-cell response activation (De Biasi et al., 2020a) and an impaired T-cell response characterized by skewing towards a Th17 phenotype (De Biasi et al., 2020b; Odak et al., 2020). Moreover, coagulation defects have been identified in critically ill patients who are prone to thrombotic complications (Helms et al., 2020; Klok et al., 2020b, 2020a). Nevertheless, the full spectrum of omics technologies has not been applied to a highly curated cohort of COVID-19 patients and controls that was established by discarding a number of key confounding factors that affect severity and death, such as older age and comorbidities, at onset.

In this study, we hypothesized that SARS-CoV-2 induces characteristic molecular changes in critical patients that can differentiate them from noncritical patients. We also hypothesized that certain host driver genes might be responsible for the development of critical illness and that those genes might represent diagnostic, prognostic, and particularly therapeutic targets. To test these hypotheses, we performed an ensemble artificial intelligence (AI)/machine learning (ML)-based multiomics study of 47 young (aged <50 years) COVID-19 patients without comorbidities admitted to the ICU and under mechanical ventilation (“critical” patients), versus matched COVID-19 patients (i.e., aged <50 years with no comorbidities) needing “only” hospitalization at a noncritical care ward (25 “noncritical” patients) and an age- and sex-matched control group of 22 healthy individuals not infected with SARS-CoV-2 (“healthy”). The multiomics approach included whole-genome sequencing (WGS), whole-blood RNA sequencing (RNA-seq), quantitative plasma and peripheral blood mononuclear cells (PBMCs) proteomics, multiplex plasma cytokine profiling and high-throughput immune cell phenotyping. These analyses were complemented by the status of anti-SARS-CoV-2 neutralizing antibodies and multitarget IgG serology as well as the measurement of neutralizing anti-type I IFN auto-antibodies in the entire cohort.

## RESULTS

### Patients’ characteristics and study design

The present study focused on patients who were hospitalized for COVID-19 at a university hospital network in northeast France (Alsace) during the first French wave of the pandemic (March-April 2020), before the routine use of corticosteroids. A total of 72 patients under 50 years of age without comorbidities were enrolled. Fifty-three of these patients were men (74%), and the median age of the patients was 40 [IQR 33; 46] years. The patients were divided into two groups: (i) a “critical” group consisting of 47 (65%) patients hospitalized at the ICU due to moderate or severe ARDS according to the Berlin criteria (Ranieri et al., 2012) with 45 requiring invasive mechanical ventilation and 2 requiring high-flow nasal oxygen and noninvasive mechanical ventilation due to acute respiratory failure and (ii) a “noncritical” group consisting of 25 patients (35%) who stayed at a noncritical care ward. In the latter group, nineteen (76%) needed low-flow supplemental oxygen. Patients who were transferred from the noncritical care ward to the ICU were considered “critical” patients. The median simplified acute physiology score (SAPS) II of the patients at the ICU was 38 [IQR 33; 47] points, and the median PaO_2_/FiO_2_ ratio of these patients was 123 [IQR 95; 168] mmHg upon admission. All the patients were discharged from the hospital or were deceased at the time of data analysis. The overall hospital- and day-28 mortality rate was 8.3% (6 patients, all in the critical group, for a mortality of 13% in this group). The characteristics of the patients in both groups are summarized in Table 1.

**Table 1.**
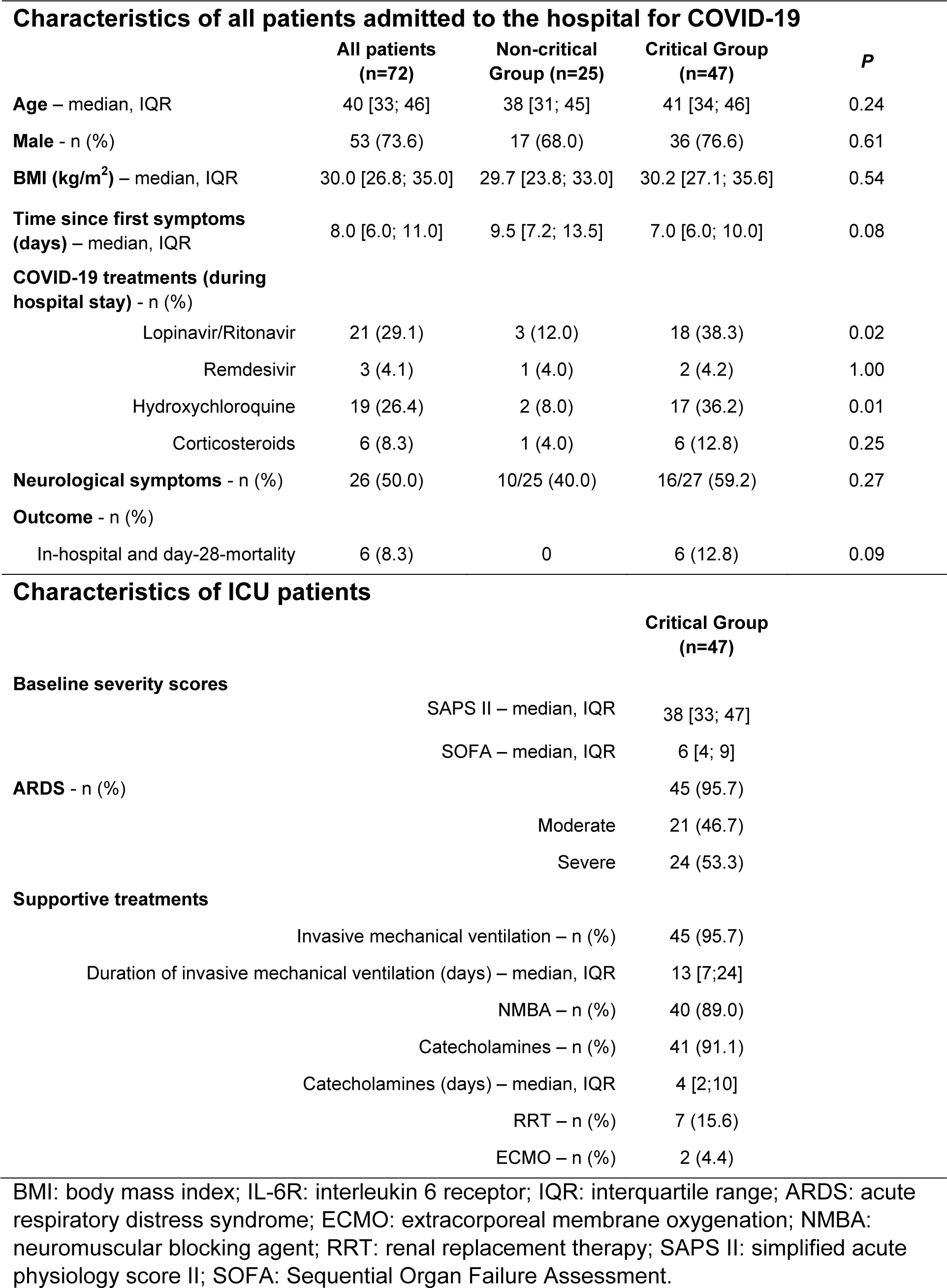
Patients description.

| **Characteristics of all patients admitted to the hospital for COVID-19** |  |  |  |  |
| --- | --- | --- | --- | --- |
|  | **All patients**<br>**(n=72)** | **Non-critical**<br>**Group (n=25)** | **Critical Group**<br>**(n=47)** | **P** |
| **Age** – median, IQR | 40 [33; 46] | 38 [31; 45] | 41 [34; 46] | 0.24 |
| **Male** - n (%) | 53 (73.6) | 17 (68.0) | 36 (76.6) | 0.61 |
| **BMI (kg/m<sup>2</sup>)** – median, IQR | 30.0 [26.8; 35.0] | 29.7 [23.8; 33.0] | 30.2 [27.1; 35.6] | 0.54 |
| **Time since first symptoms**<br>**(days)** – median, IQR | 8.0 [6.0; 11.0] | 9.5 [7.2; 13.5] | 7.0 [6.0; 10.0] | 0.08 |
| **COVID-19 treatments (during**<br>**hospital stay) - n (%)** |  |  |  |  |
| Lopinavir/Ritonavir | 21 (29.1) | 3 (12.0) | 18 (38.3) | 0.02 |
| Remdesivir | 3 (4.1) | 1 (4.0) | 2 (4.2) | 1.00 |
| Hydroxychloroquine | 19 (26.4) | 2 (8.0) | 17 (36.2) | 0.01 |
| Corticosteroids | 6 (8.3) | 1 (4.0) | 6 (12.8) | 0.25 |
| **Neurological symptoms** - n (%) | 26 (50.0) | 10/25 (40.0) | 16/27 (59.2) | 0.27 |
| **Outcome - n (%)** |  |  |  |  |
| In-hospital and day-28-mortality | 6 (8.3) | 0 | 6 (12.8) | 0.09 |
| **Characteristics of ICU patients** |  |  |  |  |
|  |  | **Critical Group**<br>**(n=47)** |  |  |
| **Baseline severity scores** |  |  |  |  |
|  | SAPS II – median, IQR | 38 [33; 47] |  |  |
|  | SOFA – median, IQR | 6 [4; 9] |  |  |
| **ARDS** - n (%) |  | 45 (95.7) |  |  |
|  | Moderate | 21 (46.7) |  |  |
|  | Severe | 24 (53.3) |  |  |
| **Supportive treatments** |  |  |  |  |
|  | Invasive mechanical ventilation – n (%) | 45 (95.7) |  |  |
|  | Duration of invasive mechanical ventilation (days) – median, IQR | 13 [7;24] |  |  |
|  | NMBA – n (%) | 40 (89.0) |  |  |
|  | Catecholamines – n (%) | 41 (91.1) |  |  |
|  | Catecholamines (days) – median, IQR | 4 [2;10] |  |  |
|  | RRT – n (%) | 7 (15.6) |  |  |
|  | ECMO – n (%) | 2 (4.4) |  |  |

BMI: body mass index; IL-6R: interleukin 6 receptor; IQR: interquartile range; ARDS: acute respiratory distress syndrome; ECMO: extracorporeal membrane oxygenation; NMBA: neuromuscular blocking agent; RRT: renal replacement therapy; SAPS II: simplified acute physiology score II; SOFA: Sequential Organ Failure Assessment.

Based on these two patient groups and an additional group of 22 healthy (SARS-Cov-2 negative) sex- and aged-matched controls, we applied a global multiomics analysis strategy to identify pathways and drivers of ARDS (Figure 1). PBMCs were analyzed by mass-cytometry (CyTOF^®^) and shotgun proteomics. Plasma samples were used for multiplex cytokine quantification and shotgun proteomics. Serum samples were used for multiplex IgG serology (Rudberg et al., 2020), detection of anti-SARS-CoV-2 neutralizing antibodies and anti-type I IFN neutralizing autoantibodies. Finally, RNA-seq and WGS were performed using whole-blood samples. Unless otherwise specified, all measures were obtained from samples that were collected at the time of hospital admission (whether at the ICU or the noncritical care ward). Validation of the identified driver genes and pathways was performed using an *ex vivo* model of SARS-CoV-2 infection and a validation cohort of 81 critical patients and 73 recovered critical patients.

**Figure 1.**
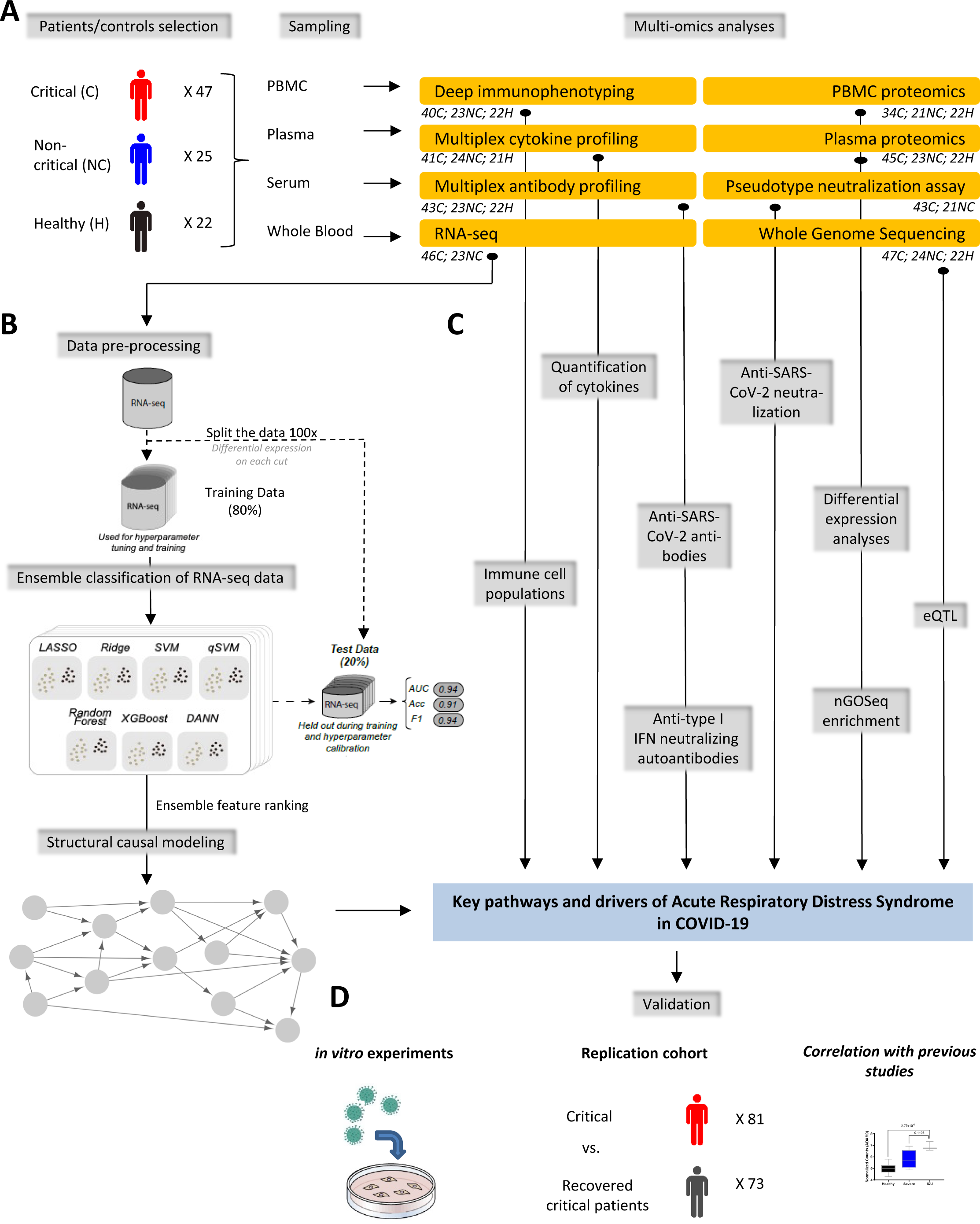
Multiomics analytical strategy. **A.** Forty-seven critical patients (C), 25 noncritical patients (NC) and 22 healthy controls (H) were enrolled in the study. PBMCs were isolated by density gradient and frozen in DMSO/FCS until utilization for Helios mass cytometry (Maxpar Direct Immune Profiling System, Fluidigm) and whole proteomics. Plasma was used for cytokine profiling (IL-17 ELISA, V-PLEX Proinflammatory Panel and S-PLEX Human IFN-α2a Kit, Mesoscale Discovery) and whole proteomics. Serum was used to measure anti-type I IFN neutralizing antibodies, anti-SARS-CoV-2 neutralizing antibodies and multi-target antiviral serology. Whole blood was used for RNA-seq (PAXgene tubes, PreAnalytiX) and whole-genome sequencing (WGS). The number of treated samples per group and per omics is indicated below each omics designation. **B.** RNA-seq pipeline based on the NC vs. C comparison. The RNA-seq data were partitioned 100 times with 80% for training and the rest for testing. For each partition of the data, feature selection was performed based on differential expression and the genes that were significantly differentially expressed in each partition of the training data were selected for both the training and corresponding test data. Classification was performed using an ensemble computational approach with seven different algorithms. After classification and verification of the quality of the results on the test dataset, an ensemble feature ranking score across six of the seven algorithms and all 100 partitions of the data was determined. The top 600 of those features were used as the input for structural causal modeling to derive a putative causal network. **C.** Cytokines and immune cells were quantified following the manufacturer’s instructions. WGS data were used for eQTL analysis together with the gene counts from the RNA-seq. Proteomics data were subjected to differential protein expression and nGOseq enrichment analyses. **D.** The key pathways and drivers resulting from the omics analyses (B and C) were validated in a replication cohort of 81 critical and 73 recovered critical patients. The differential expression of *ADAM9*, the main driver gene, was compared to publicly available bulk RNA-seq data. Finally, *ex vivo* infection experiments with SARS-CoV-2 were conducted to validate a driver gene candidate.

### Cytokines, antibodies and immune cell hallmarks of critical COVID-19

The global proinflammatory cytokine profile showed significantly increased concentrations of IFNγ, TNFα, IL-1β, IL-4, IL-6, IL-8, IL-10, and IL-12p70 in critical versus noncritical patients (Figure 2A). This “cytokine storm” (Mehta et al., 2020) was more pronounced in critical patients, as only IFNγ, TNFα and IL-10 are higher in noncritical patients as compared to healthy controls. Although the disease severity was initially associated with an RNA-seq based type I IFN signature, the absence of a significant increase in the plasma level of IFNα in critical versus noncritical patients, the decrease in the IFNα concentration during the ICU stay and the reduction in the number of plasmacytoid dendritic cells, which are the main source of IFNα, suggest that the IFN response is indeed impaired in critical patients (Figure S1) (Hadjadj et al., 2020).

**Figure 2.**
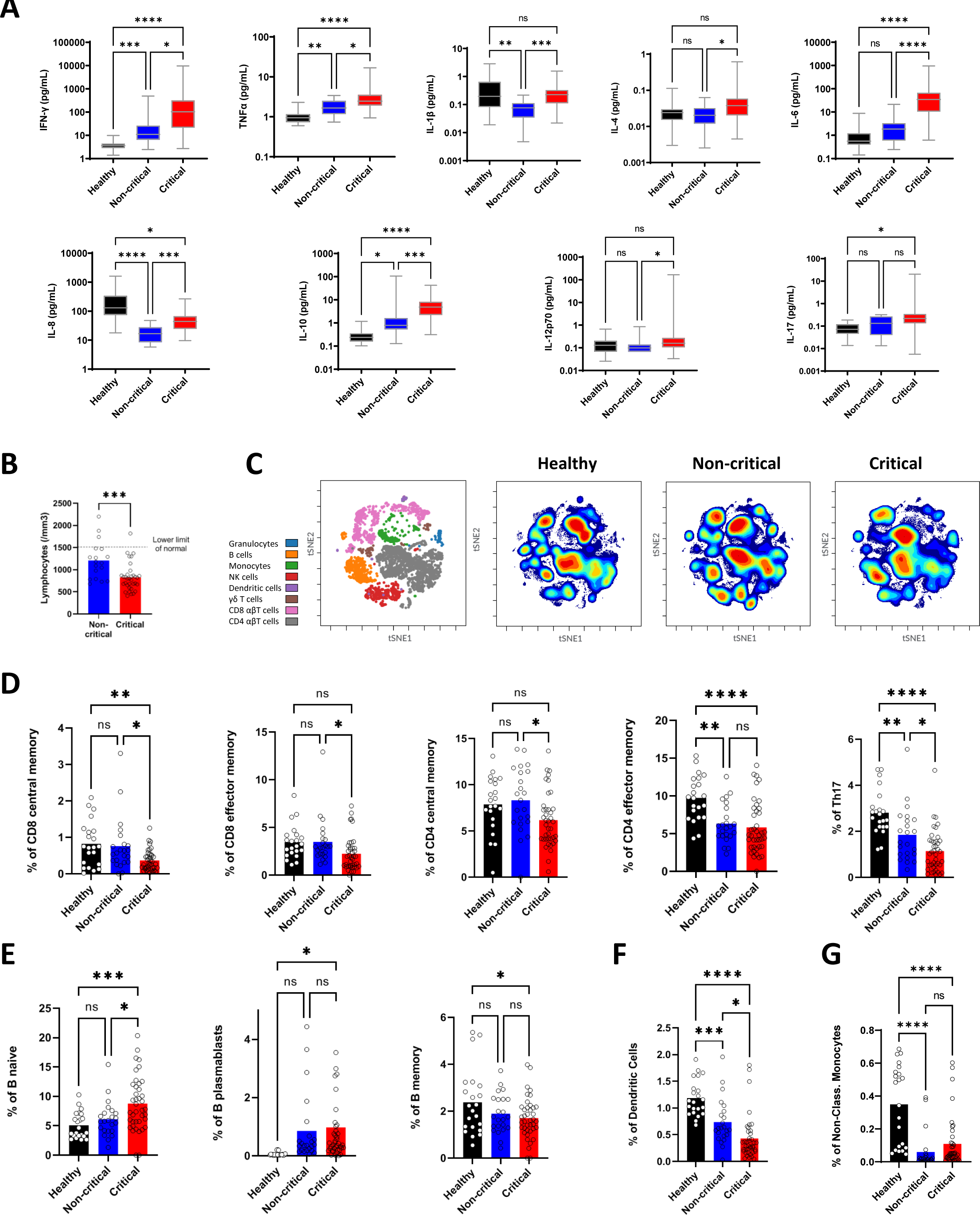
Immune profiling of healthy individuals, noncritical and critical COVID-19 patients. **A.** The levels of proinflammatory cytokines in plasma were quantified by cytokine profiling assays (V-PLEX Proinflammatory Panel and S-PLEX Human IFN-α2a Kit, Mesoscale Discovery) or ELISA (IL-17, R&D Systems). **B.** Absolute lymphocyte counts. Each dot represents a single patient. **C.** viSNE map colored according to the cell density across the three groups. Red indicates the highest density of cells. **D-G.** The proportions of modified lymphocyte subsets from COVID-19 patients and healthy controls as determined by mass cytometry. Proportions of T-cell subsets (**D**), B-cell subsets (**E**), dendritic cells (**F**) and nonclassical monocytes (**G**) are shown. The other cell subsets are presented in Figure S2. Each dot represents a single patient. In (A) and (D-G), the *P*-values were determined with the Kruskal-Wallis test, followed by Dunn’s posttest for multiple group comparisons; \**P* < 0.05, ** *P* < 0.01, *** *P* < 0.001, **** *P* < 0.0001. In (B), the *P*-value was determined by a two-tailed unpaired t-test; * *P* < 0.05, ** *P* < 0.01, *** *P* < 0.001, **** *P* < 0.0001.

At a systemic level, lymphopenia is correlated with disease severity (Guan et al., 2020; Huang et al., 2020; Mehta et al., 2020) (Figure 2B). To further characterize the immune cells, we analyzed PBMCs by mass cytometry using an immune profiling assay covering 37 cell populations. Visualization of stochastic neighbor embedding (viSNE) showed a cell population density distribution pattern that was specific to the critical group (Figure 2C). This pattern could be partly linked to the known immunosuppression phenomenon in severe patients (Hadjadj et al., 2020; Leisman et al., 2020; Remy et al., 2020), which was characterized by marked differences in the T cell compartments where memory CD4 and CD8 cells and Th17 cells were negatively correlated with disease severity (Figure 2D). The latter observation is in line with the absence of a clear association between the plasma levels of IL-17 and disease severity (Figure 2A). In contrast, the B-cell compartments of critical patients contained more naïve B cells and plasmablasts and fewer memory B cells than those of healthy controls (Figure 2E). In accordance with previous reports (De Biasi et al., 2020a), the number of plasmablasts tended to be higher in critical versus noncritical patients. Moreover, noncritical and critical patients were also characterized by lower numbers of dendritic cells and nonclassical monocytes (Figure 2F and G). The remaining cell populations are presented in Figure S2.

Altogether, the results indicate that critical illness was characterized by a proinflammatory cytokine storm and notable changes in the T, B, dendritic and monocyte cell compartments. These specific changes were independent from the extent of viral infection *per se*, as both the global anti-SARS-CoV-2 antibody levels and their neutralizing activity were not significantly different in critical versus noncritical patients (Figure S3A and B).

To complete the immunologic profile, based on findings suggesting that at least 10% of critical patients have preexisting anti-type I IFN autoantibodies (Bastard et al., 2020, 2021), we measured anti-IFNα2 and anti-IFNω neutralizing autoantibodies in patients and controls. Autoantibodies against type I IFNs were identified in two critical patients (Figure S3C) but none of the non-critical patients nor the healthy controls. Interestingly, in these two patients, the presence of autoantibodies was associated with an absence of SARS-CoV-2 neutralizing activity (Figure S3D).

### Quantitative plasma and PBMC proteomics highlight signatures of acute inflammation, myeloid activation and dysregulated blood coagulation

Quantitative nanoLC-MS/MS analysis of whole unfractionated plasma samples identified an average of 178 ± 7, 189 ± 11 and 195 ± 8 proteins in healthy individuals, noncritical and critical patients, respectively (Figure 3A). After validating the homogeneous distribution of the three groups using a multidimensional scaling plot, we performed a differential protein expression analysis to identify protein signatures that were specific to critical patients (Figure 3B and C). In line with previous studies (Chen et al., 2020b; Silvin et al., 2020), the antimicrobial calprotectin (heterodimer of S100A8 and S100A9) was among the top differentially expressed proteins (DEPs) in critical versus noncritical patients, which confirms that calprotectin is a robust marker for disease severity (Figure 3D). Our data also showed dysregulation of multiple apolipoproteins including APOA1, APOA2, APOA4, APOM, APOD, APOC1 and APOL1 (Figure 3C and E). Most of these proteins were associated with macrophage functions and were downregulated in critical patients. Acute-phase proteins (CRP, CPN1, CPN2, C6, CFB, ORM1, ORM2, SERPINA3, and SAA1) were strongly upregulated in critical patients (Figure 3C and E). These findings are consistent with previous studies showing that acute inflammation and excessive immune cell infiltration are associated with disease severity (Chen et al., 2020c; Guan et al., 2020; Shu et al., 2020).

**Figure 3.**
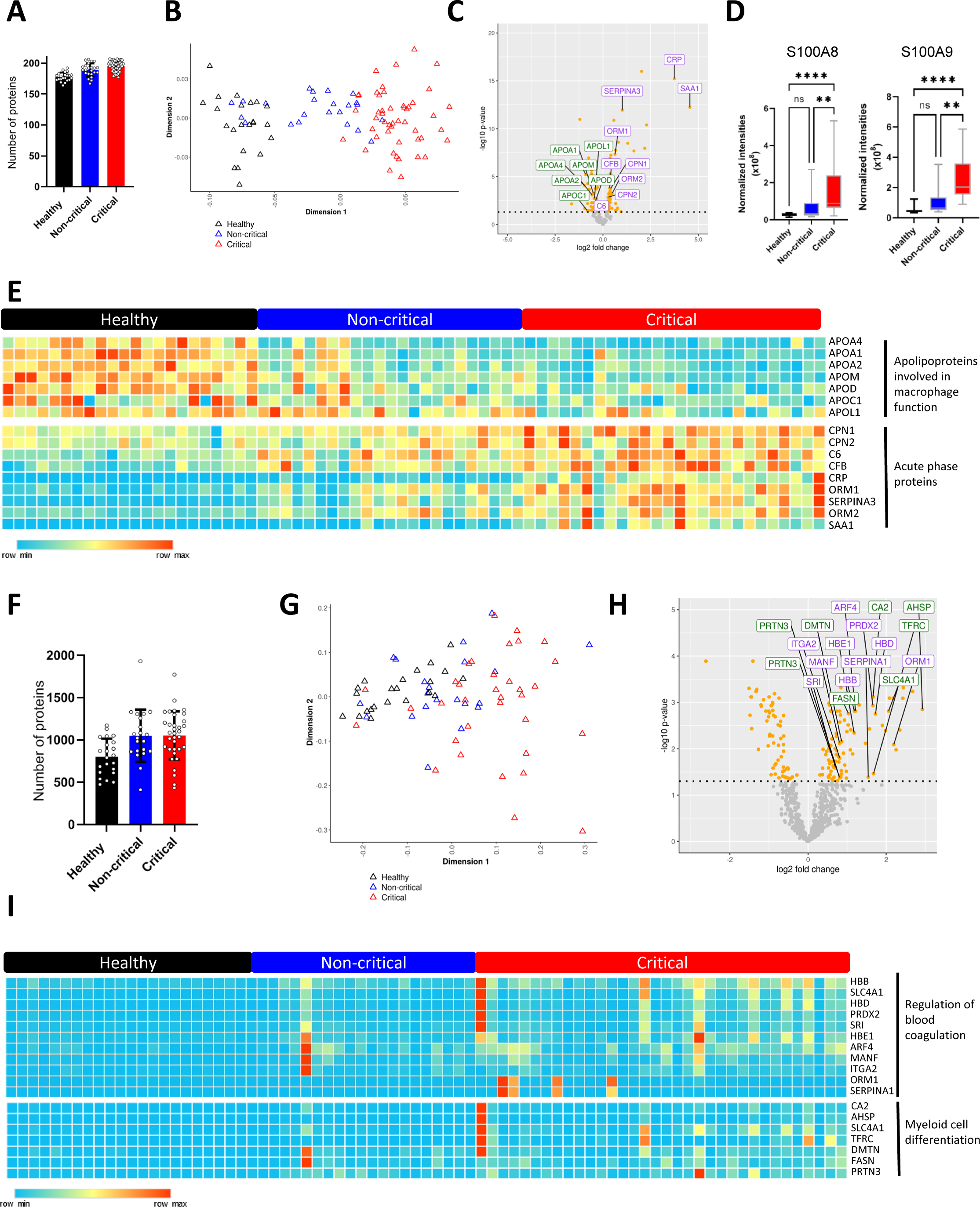
Plasma and PBMCs proteomics of healthy individuals, noncritical and critical COVID-19 patients. **A.** Total number of proteins identified in the plasma of patients and healthy controls. Each dot represents a patient. **B.** Multidimensional scaling plot of the normalized intensities of all patients/individuals in the three groups. **C.** Volcano plot representing the differentially expressed proteins (DEPs) in critical versus noncritical patients. The orange dots represent the proteins that are differentially expressed with a corrected *P*-value < 0.05. Proteins labeled in green and purple represent downregulated apolipoproteins and upregulated acute phase proteins, respectively. **D.** Normalized intensities of the proteins S100A8 and S100A9 in the three groups. *P*-values were determined with the Kruskal-Wallis test, followed by Dunn’s posttest for multiple group comparisons; \**P* < 0.05, ** *P* < 0.01, *** *P* < 0.001, **** *P* < 0.0001. **E.** Heatmap showing the expression of apolipoproteins involved in macrophage functions and acute phase proteins in the three groups. Upregulated proteins are shown in red and downregulated proteins are shown in light blue. **F.** Total number of proteins identified in PBMCs of patients and healthy controls. Each dot represents a patient. **G.** Multidimensional scaling plot of the normalized intensities of all patients/individuals in the three groups. **H.** Volcano plot representing the DEPs in critical versus noncritical patients. The orange dots represent the proteins that are differentially expressed with a corrected *P*-value < 0.05. Proteins labeled in green and purple are upregulated proteins involved in the regulation of blood coagulation and myeloid cell differentiation, respectively. **I.** Heatmap showing the expression of proteins involved in the regulation of blood coagulation and myeloid cell differentiation in the three groups. Upregulated proteins are shown in red and downregulated proteins are shown in light blue.

Whole-cell lysates of PBMCs from the same groups of patients and controls were also subjected to quantitative nanoLC-MS/MS analysis. An average of 801 ± 213, 1050 ± 309 and 1052 ± 286 proteins were identified and quantified in PBMCs of healthy individuals, noncritical and critical patients, respectively (Figure 3F). Although the distribution of the three groups in the multidimensional scaling plot was less clear than that found for plasma proteins, the differential expression analysis between noncritical and critical patients showed dysregulation of blood coagulation and myeloid cell differentiation (Figure 3H-I). The latter observation involving the CA2, AHSP, SLC4A1, TFRC, DMTN, FASN, and PRTN3 proteins was in line with the plasma proteomics results evidencing dysregulation of macrophages and with other reports showing that severe COVID-19 is marked by a dysregulated myeloid cell compartment (Schulte-Schrepping et al., 2020). The profile of the blood coagulation proteins HBB, HBD, HBE1, SLC4A1, PRDX2, SRI, ARF4, MANF, ITGA2, ORM1, and SERPINA1 confirmed that severity is also associated with coagulation-associated complications that can involve either bleeding or thrombosis (Al-Samkari et al., 2020).

### Combined transcriptomics and proteomics analysis supports inflammatory pathways associated with critical disease

Consistent with the proteomics data, differential gene expression and gene set enrichment analysis of RNA-seq data from whole blood samples collected from the patients showed that regulation of the inflammatory response, myeloid cell activation and neutrophil degranulation were the main enriched pathways in critical patients with normalized enrichment scores of 2.33, 2.65 and 2.66, respectively (Figure 4A and B).

**Figure 4.**
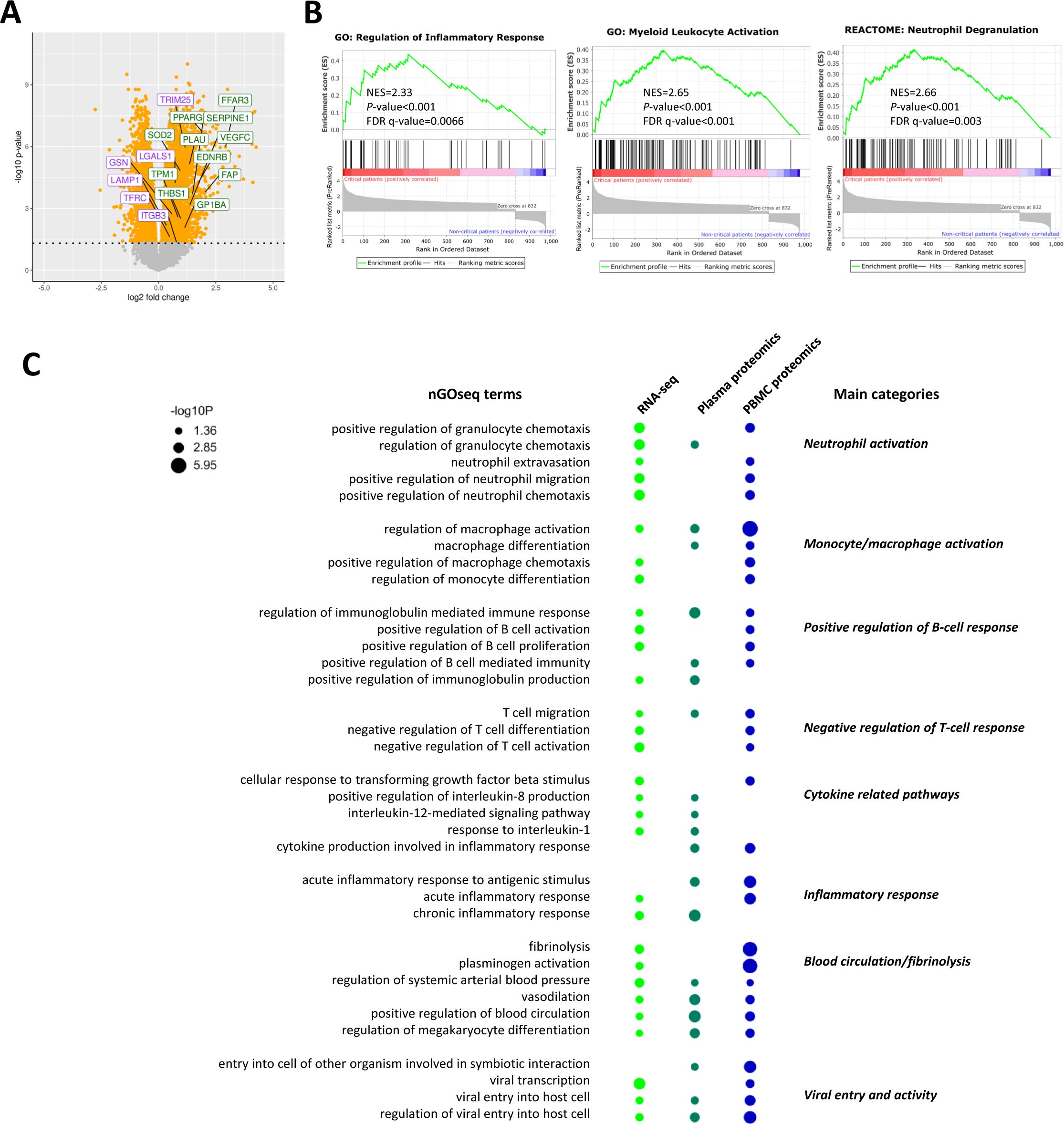
RNA-seq and combined omics analysis of critical patient-specific pathways. **A.** Volcano plot representing the differentially expressed genes in critical versus noncritical patients. The orange dots represent the genes that are differentially expressed with a corrected *P*-value < 0.05. Proteins labeled in green and purple represent upregulated genes involved in blood pressure regulation and viral entry, respectively. **B.** Gene set enrichment analysis plots showing positive enrichment of inflammatory response, myeloid leukocyte activation and neutrophil degranulation pathways. NES, normalized enrichment score. **C.** Enriched nested gene ontology (nGO) categories in critical vs. noncritical patients in RNA-seq, plasma proteomics and PBMC proteomics.

To identify enriched pathways that were supported by different omics layers, we performed nested GOSeq (nGOseq) (Yu et al., 2017) functional enrichment of the differentially expressed genes or proteins identified from the RNA-seq, plasma and PBMC proteomics data. Figure 4C shows the nGOseq terms that were found to be statistically enriched in at least two omics datasets in critical compared with non-critical patients. In line with the cytokine profiling results (Figure 2A), inflammatory signaling and the response to proinflammatory cytokine release (IL-1, IL-8 and IL-12) were supported by multiple omics datasets. As suggested by the results from immune cell profiling (Figure 2C and D) and previous studies, the B-cell response was activated, whereas the T-cell response was impaired (De Biasi et al., 2020a; Li et al., 2021a). As previously witnessed (Meizlish et al., 2021; Sánchez-Cerrillo et al., 2020; Schulte-Schrepping et al., 2020; Silvin et al., 2020), the activation of neutrophils and monocytes was confirmed by the enrichment of nine different nGOseq terms (Figure 4C). The nGOseq enrichment analysis also indicated that dysfunction of blood coagulation involves a fibrinolytic response, an observation that could, however, be linked to the anticoagulant therapy administered to most critical patients. Moreover, nGOseq terms related to viral entry and even viral transcription were strongly enriched in the three omics datasets. This result was consistent with the identification of viral gene transcripts in the RNA-seq data of eight critical patients but not in those from noncritical patients (Table S1).

### Integrated ensemble AI/ML and probabilistic programming discovers a robust gene expression signature and driver genes that differentiate critical from noncritical patients

To robustly identify a set of genes that might differentiate between noncritical and critical COVID-19 patients and could thus be related to the progression to ARDS, we adopted the pipeline depicted in Figure 1B. Briefly, we partitioned the patient blood RNA-seq data 100 times to account for sampling variation, using 80% for training and 20% for testing, and evaluated the performance of seven distinct AI/ML algorithms, including a quantum support vector machine (qSVM) to differentiate between noncritical and critical COVID-19 patients. We have previously shown that quantum annealing is a more robust classifier for relatively small patient training sets (Li et al., 2021b). The receiver operating characteristic curves (ROCs) for the 100 partitions of the patient data as well as other classification performance metrics are shown in Figure 5A and Table S2. The classification performance on the test set provided a high degree of confidence that the signals learned by the various AI/ML algorithms are generalizable.

**Figure 5.**
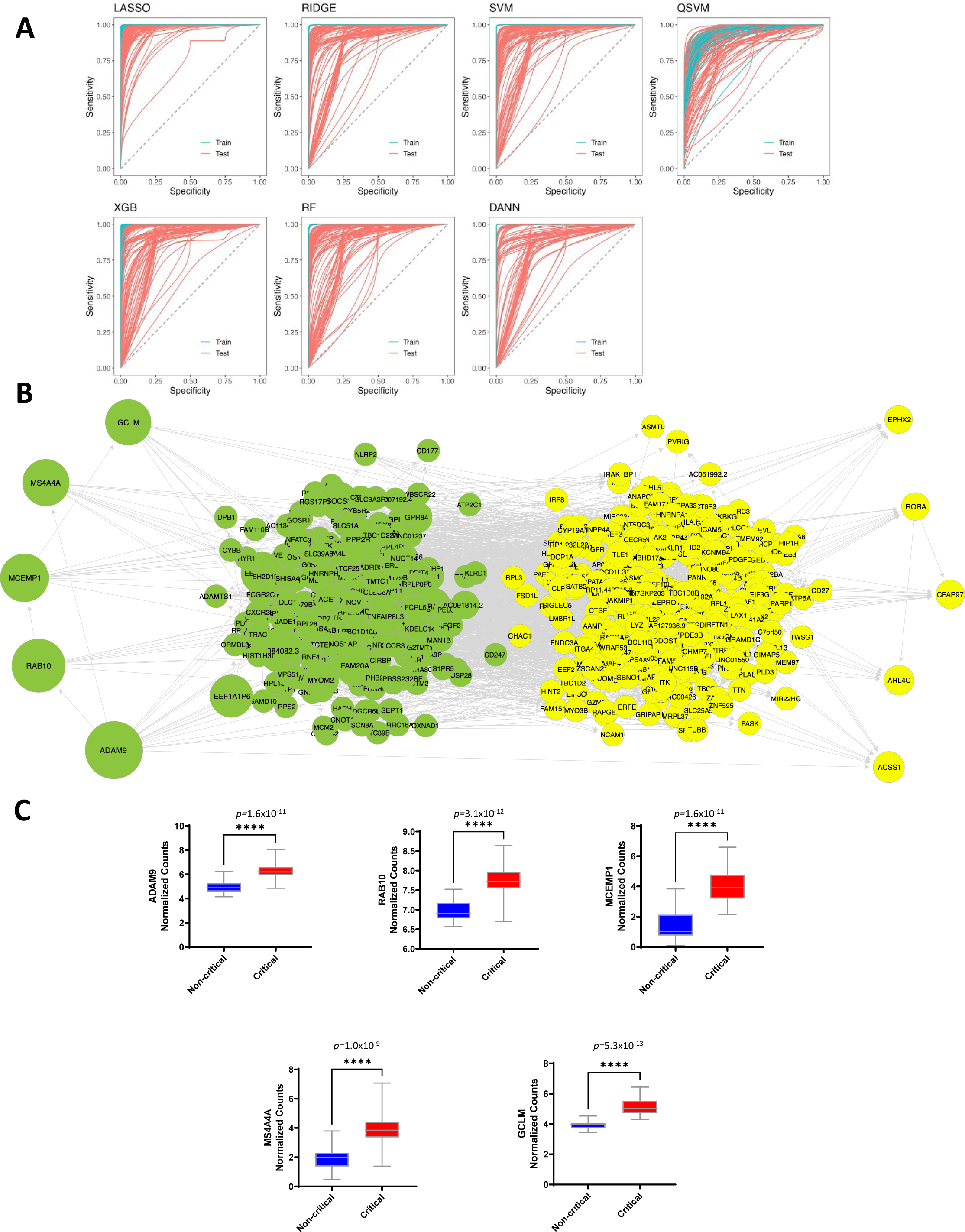
Integrated AI/ML and probabilistic programming of noncritical and critical COVID-19 patients. **A.** ROCs of the train and test sets for critical vs noncritical comparisons. All methods performed similarly. Other classification metrics are provided in Table S2. **B.** Putative network showing the flow of causal information based on the top 600 most informative genes for classifying RNA-seq data of critical versus noncritical patients. **C.** Box plots showing the normalized gene counts of the five driver genes in critical and noncritical patients. The indicated values correspond to the FDR.

After successfully classifying noncritical versus critical patients based on whole-transcriptome RNA-seq profiling, we assessed feature scores across the six distinct ML algorithms (see Methods section) and all partitions to determine an ensemble feature ranking, ignoring features from the partitions of patient data where the test AUROC was less than 0.7. Aggregating the best performing features across both the algorithm and data partitions afforded a more robust and stable set of generalizable features.

This signature represents hundreds of genes that are differentially expressed and by itself does not distinguish between driver genes of severe COVID-19 and genes that react to the disease. Therefore, we then selected the top 600 most informative genes and used them as input for structural causal modeling (SCM) to identify likely drivers of severe COVID-19 disease. Previous work has shown that SCM of RNA-seq data produces causal dependency structures, which are indicative of the signal transduction cascades that occur within cells and drive phenotypic and pathophenotypic development (Ricard et al., 2019). However, this approach works best if the gene sets are stable and consistent across six different algorithms, as shown here. The resultant SCM output is presented as a directed acyclic graph (DAG) in Figure 5B, a gene network representing the putative flow of causal information, with genes on the left predicted to have the greatest degree of influence on the entire state of the network. That is, perturbing these genes is most disruptive to the state of the network (Figure S4) and is expected to exert the greatest effect on the expression of downstream genes. The top five genes associated with the greatest degree of putative causal dependency were *ADAM9*, *RAB10*, *MCEMP1*, *MS4A4A* and *GCLM*, and all five of these genes were significantly upregulated in critical patients (Figure 5C).

The usefulness of the 600 genes identified in this first group of patients was then evaluated in a second patient cohort consisting of critical COVID-19 patients sampled at the time of entry into the ICU and recovered critical patients sampled at three months after discharge from the ICU. The top 600 genes from the first patient cohort were able to significantly differentiate between critical and recovered patients (Figure S5A and B); classification performance when training on the differentially expressed genes between critical and recovered patients was nearly the same (not shown), which indicated the high degree of generalizability of this gene signature. Moreover, the five driver genes identified in patient cohort 1 were also upregulated in critical patients in this second patient cohort (Figure S5C).

### ADAM9 is a major driver of ARDS in critical COVID-19 patients

Among the five driver genes identified by structural causal modeling, we primarily focused on experimentally determining the role of *ADAM9* (a disintegrin and a metalloprotease 9) in COVID-19 etiology because (i) it was the gene with the greatest degree of causal influence in the SCM DAG, (ii) it was the only driver gene that was previously shown to interact with SARS-CoV-2 through a global interactomics approach (Gordon et al., 2020a, 2020b) and (iii) it is an entry factor for another RNA virus, the encephalomyocarditis virus (Bazzone et al., 2019). ADAM9 is a metalloprotease with various functions that are mediated either by its disintegrin domain for adhesion or by its metalloprotease domain for the shedding of a large range of cell surface proteins (Chou et al., 2020). The *ADAM9* gene encodes two isoforms, which encode membrane-bound and secreted proteins. Although neither isoform could be detected using our proteomics approach, *ADAM9* was upregulated at the RNA level, and the secreted form was found at a higher concentration in the serum of critical versus noncritical patients (Figure 6A and B). The transcriptional upregulation of *ADAM9* was also found to be associated with disease severity in a previously published bulk RNA-seq dataset (Figure S6) (Arunachalam et al., 2020). To assess a potentially increased metalloprotease activity in the critical group, we quantified the soluble form of the MICA protein (Carapito and Bahram, 2015), which is known to be cleaved by ADAM9 (Kohga et al., 2010) by ELISA. The concentration of soluble MICA was indeed significantly higher in the plasma of critical patients as compared to noncritical patients and healthy controls (Figure 6C). A global eQTL analysis using WGS and RNA-seq data identified eight SNPs associated with three of the top five putative driver genes with genome-wide significance (Table S3). Among these SNPs, rs7840270 is localized just 0.3 kb upstream of the *ADAM9* gene and an eQTL for blood expression reported in GTEX (Figure 6D). In accordance with the observed upregulation of the gene, the higher expressing allele C was more frequent in critical than in noncritical patients (71.3% vs. 50%, OR=2.48, *P*=0.017).

**Figure 6.**
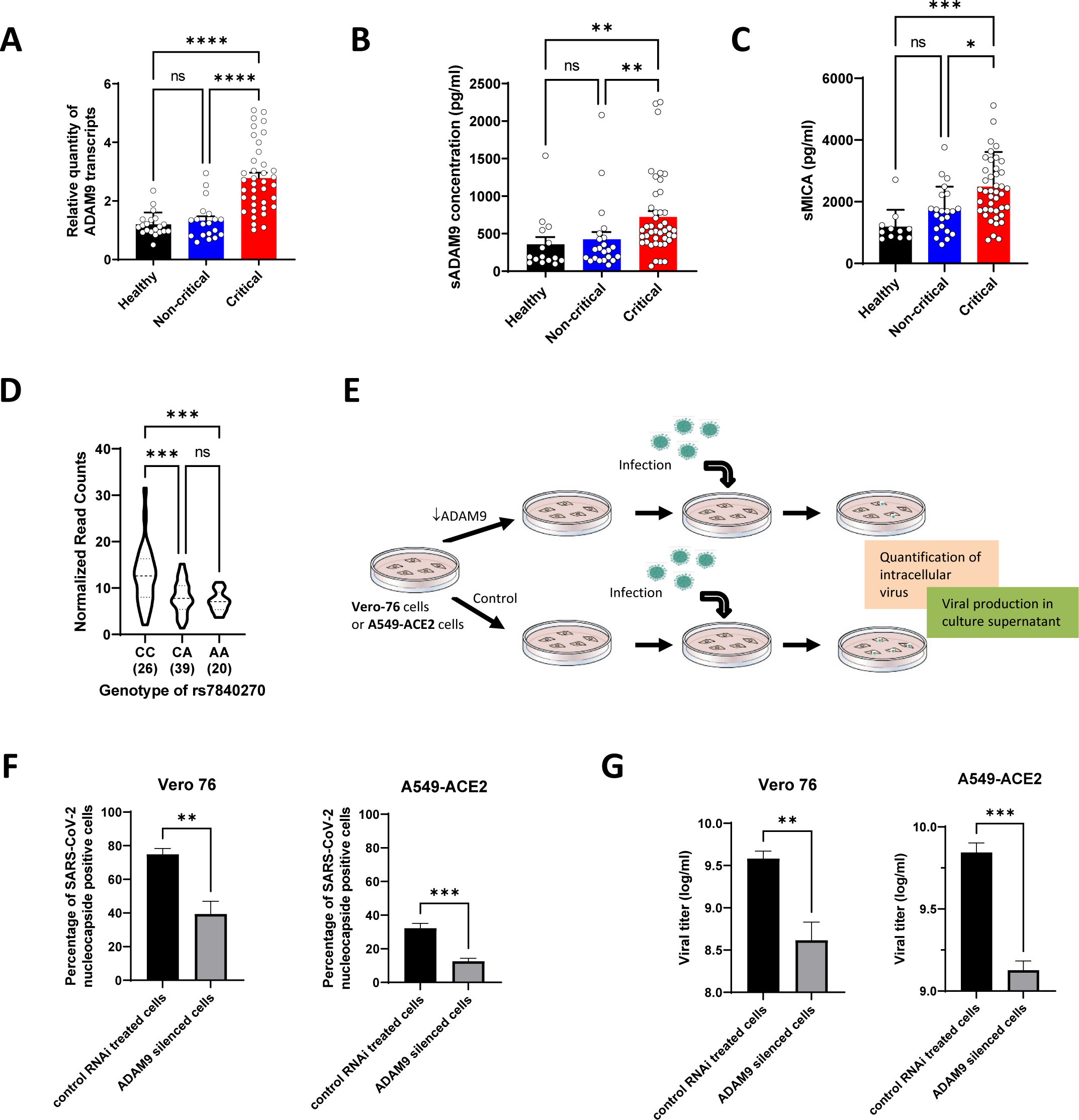
Validation of ADAM9 as a key driver of viral infection and replication. **A.** Quantitative RT-PCR confirmation of the differential expression of ADAM9 in noncritical vs. critical patients and in healthy controls. **B.** Soluble ADAM9 (sADAM9) concentration in serum of healthy controls, noncritical and critical patients determined by ELISA. **C.** Soluble MICA concentration (sMICA) in serum of healthy controls, noncritical and critical patients determined by ELISA. **D.** Expression of ADAM9 according to the genotype of the eQTL rs7840270. **E.** Experimental approach for assessing viral uptake and viral replication in silenced Vero 76 or A549-ACE2 cells. **F.** Flow cytometry-based intracellular nucleocapsid staining in control and ADAM9-silenced Vero 76 and A549-ACE2 cells. **G.** Quantitative RT-PCR of SARS-CoV-2 in culture supernatant after the silencing of *ADAM9* in Vero 76 or A549-ACE2 cells. The results from probe N1 are shown. In (A) and (F-G), the *P*-values were determined from a two-tailed unpaired t-test; * *P* < 0.05, ** *P* < 0.01, *** *P* < 0.001, **** *P* < 0.0001. In (B-D), the *P*-values were determined with the Kruskal-Wallis test followed by Dunn’s posttest for multiple group comparisons; * *P* < 0.05, ** *P* < 0.01, *** *P* < 0.001, **** *P* < 0.0001.

To assess the role of ADAM9 in viral infection, we set up an *ex vivo* assay in which *ADAM9* was silenced by siRNA in Vero 76 or A549-ACE2 (Buchrieser et al., 2020) cells and subsequently infected the cells with SARS-CoV-2. Viral replication was monitored by flow cytometry quantification of the intracellular nucleocapsid protein and by quantitative viral RT-PCR of the culture supernatant (Figure 6E). The average silencing efficiency reached 66% in Vero 76 cells and 93% in A549-ACE2 cells (Figure S7). In both cell lines, the amount of intracellular virus and the quantity of released virus were significantly lower when *ADAM9* was silenced as compared to the control condition that was treated with a control siRNA (Figure 6F and G). Our results collectively demonstrate that *ADAM9* is an *in vivo* upregulated driver in critical patients, shows higher global proteolytic activity in these patients and facilitates viral infection and replication.

## DISCUSSION

A number of studies have detailed the molecular and cellular modifications associated with COVID-19 disease severity (Arunachalam et al., 2020; Chua et al., 2020; Hadjadj et al., 2020; Lucas et al., 2020; Messner et al., 2020; Schulte-Schrepping et al., 2020; Shen et al., 2020; Shu et al., 2020; Silvin et al., 2020; Su et al., 2020; Wei et al., 2020; Zhou et al., 2020), yet very few studies have targeted a young population with no comorbidities to reduce confounders that may also drive severity and mortality, and these confounders were limited to epidemiology and/or standard bioclinical parameters such as CRP, D-dimers or SOFA scores (e.g., Ioannidis et al., 2020; Li et al., 2020; Wang et al., 2020). A comprehensive understanding of the immune responses to SARS-CoV-2 infection is fundamental to an explanation as of why some young patients without comorbidities progress to critical illness whereas others do not, a phenomenon that has been exacerbated with new viral variants in current epidemic waves across the globe (Davies et al., 2021; http://www.utisbrasileiras.com.br). In particular, knowledge of the molecular drivers of critical COVID-19 is urgently needed to identify predictive biomarkers and more efficient therapeutic targets that function through drivers of severe COVID-19 rather than to downstream or secondary events (Hermine et al., 2021; Mariette et al., 2021; Rubin et al., 2021).

Here, we used a multiomics strategy associated with integrated AI/ML and probabilistic programming methods to identify pathways and signatures that can differentiate critical from noncritical patients in a population of patients younger than 50 years without comorbidities. This *in silico* strategy provided a detailed view of the systemic immune response that was globally in accordance with previously published data. The thrust of our work, however, was to define a consistent transcriptomic signature that can robustly differentiate critical from noncritical patients, as shown by the classification performance metrics assessed in this study (Figure 5A and Table S2). Most significantly, this signature can be generalized because the same classification performance was shown to perform equally well in a replication cohort composed of 81 critically ill patients and 73 recovered critical patients (Figure S5).

Using the top 600 gene expression features of the signature as the input for structural causal modeling, we derived a causal network that uncovered five putative driver genes: *RAB10*, *MCEMP1*, *MS4A4A*, *GCLM* and *ADAM9*. RAB10 (Ras-related protein Rab-10) is a small GTPase that regulates macropinocytosis in phagocytes (Liu et al., 2020), which is a mechanism that has been suggested to be involved in the entry of SARS-CoV-2 into respiratory epithelial cells (Glebov, 2020). MCEMP1 (mast cell expressed membrane protein 1) is a membrane protein specifically associated with lung mast cells, and decreasing the expression of this protein has been shown to reduce inflammation in septic mice (Li et al., 2005; Xie et al., 2020). MS4A4A (a member of the membrane-spanning, four domain family, subfamily A) is a surface marker for M2 macrophages that mediates immune responses in pathogen clearance (Sanyal et al., 2017) and regulates arginase 1 induction during macrophage polarization and lung inflammation in mice (Sui and Zeng, 2020). GCLM (glutamate-cysteine ligase modifier subunit) is the first rate-limiting enzyme of glutathione synthesis and has been linked to severe COVID-19 (Sui and Zeng, 2020). Although these four genes are all good candidates that can at least partially explain the severity of the disease, we focused our functional validations on ADAM9, which represented, from an *in silico* standpoint, the most promising driver gene. The confirmed upregulation of ADAM9 at the RNA and protein levels in critical patients, the increased metalloprotease activity in these same patients and our *ex vivo* validation of its effect on viral uptake/replication are indeed strong arguments supporting the targeting of this protein as a potential therapeutic strategy for the treatment or prevention of severe COVID-19. ADAM9 appears to dramatically affect viral uptake. The inhibition of this presumed mechanism of action of ADAM9 might represent a novel means for the treatment SARS-CoV-2 and/or other viral illnesses. Moreover, therapies that block viral uptake rather than host receptor binding are more likely to be variant-independent, a known virological behavior which might - at least partially – derail current vaccination efforts (Dejnirattisai et al., 2021).

Due to its implication in tumor progression and metastasis, ADAM9 is currently being tested as a target of antibody-drug-conjugate therapy for solid tumors (Hicks et al., 2019). A repurposing strategy using ADAM9-blocking antibodies for the treatment of critical COVID-19 patients could therefore be envisioned. Alternatively, other therapeutic agents to reduce the ADAM9 levels or activity could be pursued.

In conclusion, this study presents a detailed multiomics investigation of a well-characterized cohort of young, previously healthy, critical COVID-19 patient series compared with noncritical patients and healthy controls. In addition to uncovering a landscape of molecular changes in the blood of critical patients, we applied a data-driven ensemble AI/ML strategy, which was independent of prior biological knowledge and thus significantly minimized possible annotation biases, to gain novel insights into COVID-19 pathogenesis and to provide potential candidate diagnostic, prognostic and especially much needed therapeutic targets that might be helpful in the ongoing battle against the COVID-19 pandemic.

## MATERIALS AND METHODS

### Patients

In March and April 2020, patients aged less than 50 years, who had no comorbidities (of note, obesity alone was not considered an exclusion criterion) and were admitted for COVID-19 to the infectious disease unit (hereafter designated noncritical care ward) or to the designated ICUs at the university hospital network in northeast France (Alsace) were investigated within the framework of the present study. Follow-up was performed until hospital discharge. SARS-CoV-2 infection was confirmed in all the patients by a quantitative real-time reverse transcriptase PCR tests for COVID-19 nucleic acid of nasopharyngeal swabs (Institut Pasteur, 2021). The ethics committee of Strasbourg University Hospitals approved the study (COVID-HUS, reference CE: 2020-34). Written informed consent was obtained from all the patients. The demographic characteristics, medical history, and symptoms were reported. Three groups were considered: (1) the “critical group” which included 47 patients admitted to the ICU, (2) the “noncritical group”, which was composed of 25 hospitalized patients at the noncritical care ward, and (3) the “healthy control group”, which included 22 healthy age- and sex-matched blood donors aged less than 50 years. A replication cohort composed of 81 critical patients and 73 recovered critical patients from one of the ICU departments of Strasbourg University hospitals was used to validate our molecular findings.

### Sampling

Venipunctures were performed upon admission to the ICU or medical ward within the framework of routine diagnostic procedures. A subset of ICU patients (73%) were sampled every 4-8 days posthospitalization until discharge or death. Patient blood was collected into BD Vacutainer tubes with heparin (for plasma and PBMCs), EDTA (for DNA) or without additive (for serum) and into PAXgene^®^ Blood RNA tubes (Becton, Dickinson and Company, Franklin Lakes, NJ, USA). Blood from healthy donors was sampled in BD Vacutainer tubes with heparin, with EDTA or without additive. Plasma and serum fractions were collected after centrifugation at 900 x g at room temperature for 10 min, aliquoted, and stored at -80°C until use. PBMCs were prepared within 24 h by Ficoll density gradient centrifugation. Aliquots of 1 x 10^6^ dry cell pellets were frozen at -80°C until use for proteomics. Aliquots of a minimum of 5 x 10^6^ cells were frozen at -80°C in 90% fetal calf serum (FCS)/10% dimethyl sulfoxide (DMSO). The EDTA and PAXgene^®^ tubes were stored at -80°C until use for DNA and RNA extraction, respectively.

### Cytokine profiling

The plasma samples were analyzed using the V-PLEX Proinflammatory Panel 1 Human Kit (IL-6, IL-8, IL-10, TNF-α, IL-12p70, IL-1β, IL-2, IL-4 and IFN-γ) and the S-PLEX Human IFN-α2a Kit following the manufacturer’s instructions (Mesoscale Discovery, Gaithersburg, MD, USA). The plasma was used undiluted for the S-PLEX Human IFN-α2a Kit and diluted 2-fold for use with the V-PLEX Proinflammatory Panel 1. The MSD plates were analyzed with an MS2400 imager (Mesoscale Discovery, Gaithersburg, MD, USA). Soluble IL-17 in undiluted serum was quantified by Quantikine^®^ HS ELISA (Human IL-17 Immunoassay) following the manufacturer’s instructions (R&D Systems, Minneapolis, MN, USA). All standards and samples were measured in duplicate.

### Immune phenotyping by mass cytometry

PBMCs were thawed rapidly, washed twice with 10 volumes of RPMI (Roswell Park Memorial Institute) medium (Thermo Fisher Scientific, Waltham, MA, USA) and centrifuged for 7 min at 300 x g at room temperature between each washing step. Cells were then treated with 250 U of DNase (Thermo Fisher Scientific, Waltham, MA, USA) in 10 volumes of RPMI medium for 30 min at 37°C in the presence of 5% CO_2_. During this step, the viability and the number of the cells were determined with Trypan Blue (Thermo Fisher Scientific, Waltham, MA, USA) and Türk’s solution (Merck Millipore, Burlington, MA, USA), respectively. After the elimination of DNase by centrifugation for 7 min at 300 x g at room temperature, a total of 3 x 10^6^ cells were used for immunostaining with the Maxpar^®^ Direct Immune Profiling Assay kit (Fluidigm, San Francisco, CA, USA), following the manufacturer’s instructions, except that we used 32% paraformaldehyde (Electron Microscopy Sciences, Hatfield, PA, USA). A red blood cell lysis step was included after the immunostaining following the manufacturer’s instructions. The prepared cells were stored at -80°C until use for acquisition with a Helios mass cytometer system (Fluidigm, San Francisco, CA, USA). An average of 600000 events were acquired per sample. The mass cytometry standard files produced with the Helios instrument were analyzed using Maxpar^®^ Pathsetter software v.2.0.45 that was modified for live/dead parameters: the tallest peak was selected instead of the closest peak for the identification and quantification of the cell populations. The FCS files from each group (healthy, critical, non-critical) were then concatenated using CyTOF^®^ software v.7.0.8493.0 for viSNE analysis (Cytobank Inc, Mountain View, CA, USA). A total of 300000 events were used for the viSNE map that was generated with the following parameters: iterations (1000), perplexity (30) and theta (0.5). viSNE maps are presented as the means of all samples in each group.

### Plasma proteomics analysis

#### Sample preparation

Samples were prepared using the PreOmics iST Kit (PreOmics GmbH, Martinsried, Germany) according to the manufacturer’s protocol. Two microliters of plasma was mixed with 50 µl of Lyse buffer and heated at 95°C for 10 min at 1000 rpm. The protein concentration was determined using the Bradford assay (Bio-Rad, Hercules, CA, USA) according to the manufacturer’s instructions. The samples were transferred to 96-well-plate cartridges, and 50 µl of resuspended Digest solution was added. The samples were then heated at 37°C for 2 h, and 100 µl of Stop buffer was added. The samples were centrifuged to retain the peptides on the cartridge and washed twice with “Wash 1” and “Wash 2” buffers. The peptides were then eluted twice with Elute buffer before evaporation under vacuum. The peptides were then resuspended using the “LC-load” solution containing iRT peptides (Biognosys, Schlieren, Switzerland), and the samples were rapidly sonicated before being injected into the nanoLC-MS/MS system.

#### NanoLC-MS/MS analysis

The NanoLC-MS/MS analyses were performed with a nanoAcquity Ultra-Performance LC^®^ (UPLC^®^) device (Waters Corporation, Milford, MA, USA) coupled to a Q-Exactive^TM^ Plus mass spectrometer (Thermo Fisher Scientific, Waltham, MA, USA). Peptide separation was performed on an ACQUITY UPLC BEH130 C18 column (250 mm × 75 μm with 1.7-μm-diameter particles) and a Symmetry C18 precolumn (20 mm × 180 μm with 5-μm-diameter particles, Waters). The solvent system consisted of 0.1% FA in water (solvent A) and 0.1% FA in ACN (solvent B). The samples (equivalent to 500 ng of proteins) were loaded into the enrichment column over 3 min at 5 μl/min with 99% solvent A and 1% solvent B. The peptides were eluted at 400 nl/min using the following gradient of solvent B: from 1 to 35% over 60 min and from 35 to 90% over 1 min. The 90 samples were injected in randomized order. The MS capillary voltage was set to 2.1 kV at 250°C. The system was operated in the data dependent acquisition mode with automatic switching between MS (mass range 300–1800 m/z with R = 70000, automatic gain control (AGC) fixed to 3 x 10^6^ ions, and maximum injection time of 50 ms) and MS/MS (mass range of 200–2000 m/z with R = 17,500, AGC fixed at 1 x 10^5^ and maximal injection time of 100 ms) modes. The ten most abundant ions were selected on each MS spectrum for further isolation and higher energy collision dissociation fragmentation, excluding unassigned and monocharged ions. The dynamic exclusion time was set to 60s. A sample pool comprising equal amounts of all protein extracts was constituted and regularly injected during the course of the experiment as an additional quality control.

#### Data analysis

The raw data obtained from each sample (45 critical patients, 23 non-critical patients, and 22 healthy controls) were processed using MaxQuant (version 1.6.14). Peaks were assigned using the Andromeda search engine with trypsin/P specificity. A database containing all human entries was extracted from the UniProtKB-SwissProt database (May 11 2020, 20410 entries). The minimal peptide length required was seven amino acids, and a maximum of one missed cleavage was allowed. Methionine oxidation and acetylation of the proteins’ N-termini were set as variable modifications, and acetylated and modified methionine-containing peptides, as well as their unmodified counterparts, were excluded from the protein quantification step. Cysteine carbamidomethylation was set as a fixed modification. The “match between runs” option was enabled. The maximum false discovery rate was set to 1% at the peptide and protein levels with the use of a decoy strategy. The normalized label-free quantification (LFQ) intensities were extracted from the ProteinGroups.txt file after the removal of nonhuman and keratin contaminants, as well as reverse and proteins only identified by site. This resulted in 336 quantified proteins. Complete datasets have been deposited in the ProteomeXchange Consortium database with the identifier PXD025265 (Deutsch et al., 2017).

#### Differential protein expression analysis

The LFQ values from MaxQuant were used for differential protein expression analysis. For each pairwise comparison, the proteins expressed in at least 80% of the samples in either group were retained. Variance stabilization normalization (Vsn) was performed using the justvsn function from the vsn R package (Huber et al., 2002). Missing values were imputed using the random forest approach (Kokla et al., 2019). This process resulted in 161 proteins. Differential protein expression analysis was performed using the limma bioconductor package in R (Ritchie et al., 2015). Significant differentially expressed proteins were determined based on an adjusted *P*-value cutoff of 0.05 using the Benjamini-Hochberg method.

### PBMC proteomics analysis

Samples were prepared using the PreOmics’ iST Kit (PreOmics GmbH, Martinsried, Germany) according to the manufacturer’s protocol. Briefly, PBMC pellets were resuspended in 50 µl of Lyse buffer and heated at 95°C for 10 min at 1000 rpm before being sonicated for 10 min on ice. The protein concentration of the extract was determined using the Pierce™ BCA Protein Assay Kit (Thermo Fisher, Waltham, MA, USA). The samples were transferred to 96-well plate cartridges and 50 µl of resuspended Digest solution was added. The samples were then heated at 37°C for 2 h and 100 µl of Stop buffer was added. The samples were centrifuged to retain the peptides on the cartridge and washed twice with “Wash 1” and “Wash 2” buffers. The peptides were then eluted twice with Elute buffer before evaporation under vacuum. Finally, the peptides were resuspended using the “LC-load” solution containing iRT peptides (Biognosys, Schlieren, Switzerland), and the samples were rapidly sonicated before being injected into the nanoLC-MS/MS system.

#### NanoLC-MS/MS analysis

NanoLC-MS/MS analyses were performed with a nanoAcquity UPLC device (Waters Corporation, Milford, MA, USA) coupled to a Q-Exactive HF-X mass spectrometer (Thermo Fisher Scientific, Waltham, MA, USA). Peptide separation was performed on an Acquity UPLC BEH130 C18 column (250 mm × 75 μm with 1.7-μm-diameter particles) and a Symmetry C18 precolumn (20 mm × 180 μm with 5-μm-diameter particles, Waters). The solvent system consisted of 0.1% formic acid (FA) in water (solvent A) and 0.1% FA in acetonitrile (ACN) (solvent B). The samples (equivalent to 414 ng of proteins) were loaded into the enrichment column over 3 min at 5 μl/min with 99% solvent A and 1% solvent B. The peptides were eluted at 400 nl/min using the following gradient of solvent B: from 2 to 25% over 53 min, from 25 to 40% over 10 min and 40 to 90% over 2 min. The 77 samples were injected using a randomized injection sequence. The MS capillary voltage was set to 1.9 kV at 250°C. The system was operated in data dependent acquisition mode with automatic switching between MS (mass range of 300-1800 m/z with R = 60000, automatic gain control (AGC) fixed to 3 x 10^6^ ions and maximum injection time of 50ms) and MS/MS (mass range of 200–2000 m/z with R = 15000, AGC fixed at 1 x 10^5^ and maximal injection time of 100 ms) modes. The ten most abundant ions were selected on each MS spectrum for further isolation and higher energy collision dissociation fragmentation with the exclusion of unassigned and monocharged ions. The dynamic exclusion time was set to 60 s.

#### Data analysis

The raw data obtained from each sample (34 critical patients, 21 non-critical patients and 22 healthy controls) were processed using MaxQuant (version 1.6.14). Peaks were assigned using the Andromeda search engine with trypsin/P specificity. A combined human and bovine database (because of potential traces of fetal calf serum in the samples) was extracted from UniProtKB-SwissProt (8 September 2020, 26,413 entries). The minimal peptide length required was seven amino acids and a maximum of one missed cleavage was allowed. Methionine oxidation and acetylation of the proteins’ N-termini were set as variable modifications, and acetylated and modified methionine-containing peptides, as well as their unmodified counterparts, were excluded from protein quantification. Cysteine carbamidomethylation was set as a fixed modification. The “match between runs” option was enabled. The maximum false discovery rate was set to 1% at the peptide and protein levels with the use of a decoy strategy. Only peptides unique to human entries were retained and their LFQ intensities were summed to derive the protein intensities. This process resulted in 2,196 quantified proteins. Complete datasets have been deposited in the ProteomeXchange Consortium database with the identifier PXD 025265 (Deutsch et al., 2017).

#### Differential protein expression analysis

Summed peptides normalized label-free quantification (LFQ values from MaxQuant software) values were used for differential protein expression analysis. For each pairwise comparison, proteins expressed in at least 80% of the samples in either group were retained. Variance stabilization normalization (Vsn) was performed using the justvsn function from the vsn R package (Huber et al., 2002). Missing values were imputed using the random forest approach (Kokla et al., 2019). This resulted in 732 proteins. Differential protein expression analysis was performed using the limma bioconductor package in R (Ritchie et al., 2015). Significant differentially expressed proteins were determined based on an adjusted *P*-value cutoff of 0.05 using the Benjamini-Hochberg method.

### Serology analysis

The serum IgG reactivity towards three SARS-CoV-2 viral antigens was measured as previously described (Rudberg et al., 2020) by means of a multiplex antigen bead array. The three viral antigens included in our multiplex bead array comprise two representations of the Spike protein (a soluble trimeric form of the spike glycoprotein stabilized in the pre-fusion conformation expressed in HEK, and the Spike S1 domain expressed in CHO cells), and the C-terminal domain of the Nucleocapsid protein (expressed in *E.coli*). Briefly, each antigen was immobilized on the surface of uniquely color-coded magnetic beads (Bead ID; MagPlex^®^, Luminex Corporation, Austin, TX, USA), and the bead IDs pooled together to generate the bead array used to test the serum samples. Sera were incubated with the multiplex antigen array in a 384-well plate format, and the IgG reactive to the viral antigen was detected by means of a phycoerythine-conjugated anti-hIgG (H10104, Invitrogen, Thermo Fisher Scientific, Waltham, MA, USA) in a FlexMap3D instrument (Luminex Corporation, Austin, TX, USA). The cut-off for reactivity was evaluated for each antigen singularly and defined as the mean +6SD of the intensity signals of 12 negative controls. The 12 controls were carefully selected among pre-pandemic samples as representative of the background range for each single antigen included in the test. The performance of this serology assay was previously evaluated on 2090 negative samples (pre-pandemic samples collected in 2019 or earlier and including 26 samples from individuals infected by other Coronaviruses) and 331 samples from COVID-19 PCR-confirmed cases (sampled collected at least 17 days after disease onset), showing 99.7% sensitivity and 100% specificity.

### Neutralizing antibodies

Spike pseudotype neutralization assay was conducted using vesicular stomatitis virus (VSV) where glycoprotein gene (G) has been deleted and substituted in trans with SARS-CoV-2 Spike protein (D614G variant) lacking terminal eighteen amino acids of the cytoplasmic domain. The pseudotype VSV-ΔG SARS-CoV-2 S D614G is cytopathic and expresses the firefly luciferase. For the neutralization assay, VSV-ΔG SARS-CoV-2 S D614G was incubated with serial dilutions of patient sera starting at 1:40 dilution for 1 h at room temperature. A monolayer of Vero cells seeded at 60000 cells/well in black-well plates was infected with the pseudotype virus (virus only) or virus and serum mixture overnight at 37°C - 5% CO_2_. The following day, cells were lysed using passive lysis buffer (Promega, Madison, WI, USA) and luciferase activity was measured upon addition of substrate (Promega^®^ CellTiterGlo^®^, Madison, WI, USA) using Biotek plate reader and Gen5 software. Neutralization was calculated by comparing relative luciferase units of serum treated versus the virus only control. To determine neutralizing titers, the concentration of the antibody resulting in 50% neutralization (NT_50_) was determined using XLFit and graphed using Prism (GraphPad, San Diego, CA, USA).

### Autoantibodies against type I IFNs

The blocking activity of anti-IFNα and anti-IFNω autoantibodies was determined by assessing a reporter luciferase activity as previously described (Bastard et al., 2021). Briefly, HEK293T cells were transfected with the firefly luciferase plasmids under the control of human ISRE promoters in the pGL4.45 backbone, and a constitutively expressing Renilla luciferase plasmid for normalization (pRL-SV40). Cells were transfected in the presence of the X-tremeGene 9 transfection reagent (Sigma Aldrich, Saint-Louis, MI, USA) for 36 hours. Then, Dulbecco’s modified Eagle medium (DMEM, Thermo Fisher Scientific, Waltham, MA, USA) supplemented with 10% healthy control or patient serum and were either left unstimulated or were stimulated with IFNα or IFNω (10 ng/mL) for 16 hours at 37°C. Each sample was tested once. Finally, Luciferase levels were measured with the Dual-Glo reagent, according to the manufacturer’s protocol (Promega, Madison, WI, USA). Firefly luciferase values were normalized against Renilla luciferase values, and fold induction is shown relative to controls transfected with empty plasmids.

### Whole-genome sequencing (WGS)

WGS data was generated from DNA isolated from whole blood. Novaseq^TM^ 6000 (Illumina Inc., San Diego, CA, USA) machines were used for DNA sequencing to a mean 30X coverage. The raw sequencing reads from FASTQ files were aligned using Burrows-Wheeler Aligner (BWA) (Li and Durbin, 2009), and GVCF files were generated using Sentieon version 201808.03 (Kendig et al., 2019). Functional annotation of the variants was performed using Variant Effect Predictor from Ensembl (version 101). GATK version 4 (Van der Auwera et al., 2013; DePristo et al., 2011) was used for the joint genotyping process and variant quality score recalibration (VQSR). We removed one duplicate sample based on kinship (king cutoff of 0.3) and retained 24,476,739 SNPs that were given a ‘PASS’ filter status by VQSR. The analysis of the 72 samples from the critical and non-critical groups identified 15870076 variants with MAF < 5%. The first two principal components were generated using plink2 on LD-pruned variants with Hardy-Weinberg equilibrium in the controls with a *P*-value ≥ 1 × 10^−6^ and MAF > 5% and were used as covariates to correct for population stratification.

### Analysis of expression quantitative trait loci (eQTLs)

We performed local (cis-) eQTL analysis to test for associations between genetic variants and gene expression levels in 67 samples having both RNA-seq and SNP genotype data. Briefly, we used the MatrixEQTL R package (Shabalin, 2012) where we selected a linear model and a maximum distance for gene-SNP pairs of 1 × 10^6^. The top two principal components identified from the genotype principal component analysis were used as covariates to control for population stratification. We selected 304044 significant eQTLs with false discovery rate (FDR) ≤ 0.05.

### RNA sequencing (RNA-seq)

#### RNA extraction

Whole-blood RNA was extracted from PAXgene tubes with the PAXgene Blood RNA Kit following the manufacturer’s instructions (Qiagen, Hilden, Germany). A total of 69 samples, including 46 critical and 23 non-critical patients were processed. The RNA quantity and quality were assessed using the Agilent 4200 TapeStation system (for the RIN) (Agilent Technologies, Santa Clara, CA, USA) and RiboGreen^TM^ (for the concentration) (Thermo Fisher Scientific, Waltham, MA, USA). RNA sequencing libraries were generated using the TruSeq Stranded Total RNA with Ribo-Zero Globin kit (Illumina, San Diego, CA, USA) and sequenced on the Illumina NovaSeq 6000 instrument with S4 flow cells and 151-bp paired-end reads. The raw sequencing data were aligned to a reference human genome build 38 (GRCh38) using the short reads aligner STAR (Dobin et al., 2013). Quantification of gene expression was performed using RSEM (Li and Dewey, 2011) with GENCODE annotation v25 (http://www.gencodegenes.org). Raw and processed datasets have been deposited in GEO with identifier GSE172114.

#### Differential gene expression (DGE) analysis

DGE analysis was performed for two different purposes: 1) for the combined omics analysis of differentially expressed genes and proteins, and 2) as step to determine feature selection for classification in the *in silico* computational intelligence approach. For the combined omics analysis, we first removed low expressed genes for the 69 samples by removing genes with less than 1 count per million in less than 10% of the samples. We then performed DGE analysis on all 69 samples using the trimmed mean of M-values method (TMM) from the edgeR R package (Robinson and Oshlack, 2010; Robinson et al., 2010).

In our computational intelligence approach, we performed DGE analysis for each partition of the train data using a frozen TMM normalization to calculate normalization factors based only on the training data, in order to avoid data leakage. Briefly, we removed low expressed genes for the 69 samples with genes with 1 count per million in less than 10% of samples. For each partition of the training data, we calculated the normalization factors, and then selected the library that had a normalization factor closest to 1. We used this library as a reference library to normalize all the samples keeping the training normalization factors unchanged. Differentially expressed genes were identified using quasi-likelihood F-test (QLF)-adjusted *P*-values from the edgeR R package. Differentially expressed genes with FDRs less than 0.05 were used for further downstream analysis.

#### Identification of potential driver genes through structural causal modeling

To identify potential biomarkers that might differentiate patients in the noncritical group from those in the critical group, we used classification as a feature selection approach and then used the most informative features as input for structural causal modeling to identify potential driver genes. More specifically, classification was performed using the RNA-seq data by repeatedly partitioning noncritical and critical patients into 100 unique training and independent test sets representing 80% and 20% of the total data, respectively, ensuring that the proportions of noncritical and critical patients were consistent in each partition of the data. One hundred partitions of the data were used to capture the biological variation and to obtain increased statistical confidence in the results. After classification, feature scores for each method were determined and combined across all 100 partitions of the data and six of the ML algorithms, not including the deep learning algorithm. The 600 most informative features were retained for structural causal modeling. More details about the classification algorithms used, the feature ranking, and the structural causal modeling are provided below.

#### Ensemble artificial intelligence

We used seven distinct ML approaches for our classification models. The relevant hyperparameters for each method are mentioned in their respective sections. Hyperparameters were selected by using 10-fold cross-validation of the training data, and the performance was evaluated using the held-out test data.

#### Least Absolute Shrinkage and Selection Operator (LASSO), and Ridge Regression

LASSO (Tibshirani, 1996) is an L1-penalized linear regression model defined as:

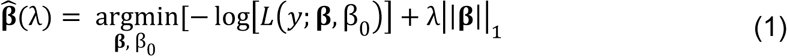

Ridge (Hoerl and Kennard, 1970; Hoerl et al., 1975) is an L2-penalized linear regression model defined as:

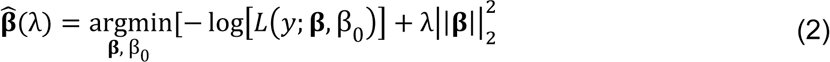

where the loss function is

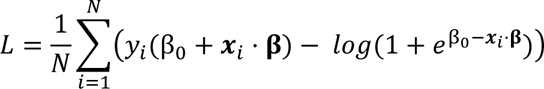

In both cases, λ > 0 is the regularization parameter that controls model complexity, β are the regression coefficients, β_0_ is the intercept term, *y* represents an indicator function f or the critical patients (i.e., *y*_i_ = 1 if the *i*-th training sample is a critical patient; otherwise *y*_*i*_ = 0), the vector *x*_*i*_ is the *i*-th training sample, and the goal of the training procedure is to determine 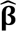 and *β*_0_, the optimal regression coefficients and the optimal intercept, that minimize the quantities defined in Eqs. (1) and (2). The predicted label is given by *y* = β_0_ + *x* ⋅ β, where a threshold of 0.5 is introduced to binarize the label for classification problems. In LASSO, the constraint placed on the norm of β (the strength of which is given by λ) causes coefficients of uninformative features to shrink to zero. This leads to a simpler model that contains only a few nonzero coefficients. We used the ‘glmnet’ function from the caret (Kuhn, 2008) R package to train all Lasso and Ridge models. For Ridge, the constraint placed on the norm of β plays a similar role in determining model complexity, except that coefficients for uninformative features do not necessarily shrink to zero.

For both Lasso and Ridge, we opted to implement the function over a custom tuning grid of λ from 2^−8^ to 2^2^. λ was selected via 10-fold cross-validation as the value that gave the minimum mean cross-validated error.

#### Support Vector Machines (SVM)

SVMs (Boser et al., 1992; Cortes and Vapnik, 1995) are a set of supervised learning models used for classification and regression analyses. The primal form of the optimization problem is:

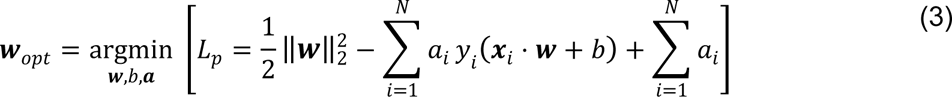

where *L*_*p*_ is the loss function in its primal form (*p* for primal), *w* represents the weights to be determined in the optimization, *x*_*i*_ is the *i*-th training sample, *y*_*i*_ is the label of the *i*-th training sample, *a*_*i*_ ≥ 0 are Lagrange multipliers, *N* is the number of training points, and *b* is the intercept term. Labels are predicted by thresholding *x*_*i*_ ⋅ *w* + *b*.

The optimization problem in its dual form is defined as

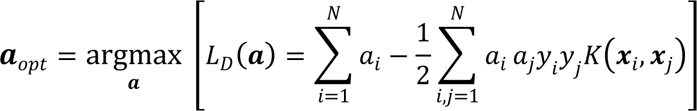

where *L*_*D*_ is the Lagrangian dual of the primal problem, *a*_*i*_ are the Lagrange multipliers, *y*_*i*_ and *x*_*i*_ are the *i*-th label and training sample, respectively, and *K*⋅,⋅) is the kernel function. Maximization takes place subject to the constraints ∑_*i*_ *a*_*i*_ *y*_*i*_ = 0 and *a*_*i*_ ≥ *C* ≥ 0, ∀*i*. Here *C* is a hyperparameter that controls the degree of misclassification of the model for nonlinear classifiers. The optimal values of *w* and *b* can be found in terms of the *a*_*i*_’s, and the label of a new data point *x* can be found by thresholding the output ∑_*i*_ *a*_*i*_ *y*_*i*_ *K*(*x*_*i*_, *x*) + *b*.

In most cases, many of the *a*_*i*_’s are zero, and evaluating predictions can be faster using the dual form. We used SVM with a linear kernel (‘svmLinear2’) (i.e., *Kx*_*i*_, *x*_*j*_) = *x*_*i*_ ⋅ *x*_*j*_, the inner product of *x*_*i*_ and *x*_*j*_) function from the caret (Kuhn, 2008) R package to train all SVM models. *C* ranged from 2^−2^ to 2^3^, and a 10-fold cross-validation was used to tune and select the hyperparameters with the best cross-validation accuracy for training the model.

#### Random Forest (RF)

RF (Breiman, 2001; Breiman et al., 1993) is an ensemble learning method for classification and regression that builds a set (or forest) of decision trees. In RF, *n* samples are selected (typically two-thirds of all the training data) with replacement from the training data *m* times, which yields *m* different decision trees. Each tree is grown by considering ‘mtry’ of the total features, and the tree is split depending on which features yield the smallest Gini impurity. In the event of multiple training samples in a terminal node of a particular tree, the predicted label is given by the mode of all the training samples in a terminal node. The final prediction for a new sample *x* is determined by taking the majority vote over all the trees in the forest. We used the ‘rf’ function from the caret (Kuhn, 2008) R package to train all RF models. Ten-fold cross-validation was used to tune the parameters for training the model. A tune grid with 44 values from 1 to 44 for ‘mtry’, the number of random variables considered for a split in each iteration during the construction of each tree, was used for the tuning model.

#### XGBoost (XGB)

XGB (Chen and Guestrin, 2016) is a distributed gradient boosting library for classification and regression by building an ensemble of decision trees. In contrast to RF, XGB uses an additive strategy to add new trees one at a time based on whether they optimize the objective function. The objective function for the *t*-th tree is

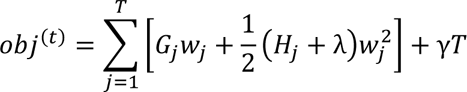

where 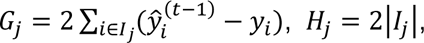, λ and γ are hyperparameters controlling model complexity, *T* is the number of leaves in the trees, *W*_*j*_ is the combined score across all the data points for the *j*-th leaf. Here, *I*_*j*_ refers to the set of indices of data points assigned to the *j*-th leaf, |*I_j_*| is the size of the set 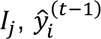 is the predicted score (without the *t*-th tree) of the *i*-th data point, and *y*_*i*_ is the actual label of the *i*-th data point. The default parameter tuning grid in R was used, and a 10-fold cross-validation was used to tune and select the hyperparameters with the best cross-validation accuracy for training the model.

#### Quantum Support Vector Machines (qSVM)

qSVM is a quantum adaptation of SVM that can be used for classification designed to be run with a quantum annealer (QA) (Willsch et al., 2020). The advantage of running the optimization problem on a QA is that, since the QA samples from the quantum distribution, it retains both the lowest energy solution and some of the next lowest-energy solutions (Albash and Lidar, 2018). Because of the suboptimal solutions, we expect qSVM to perform worse on the training data than classical SVM (which only includes the optimal solution). However, suboptimal solutions can capture different aspects of the training data and generate different decision boundaries. As such, a suitable combination of the suboptimal solutions in qSVM might outperform cSVM on the test data.

The objective function is the same as for classical SVM up to a change in sign, i.e.,

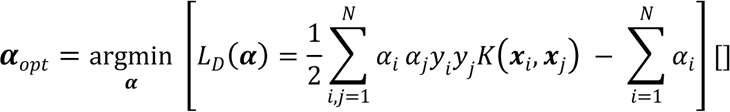

subject to the constraints 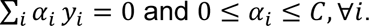

qSVM was run on physical quantum annealers manufactured by D-Wave Inc. (Johnson et al., 2011). The D-Wave “Advantage” device used in this work had 5436 qubits with 15 couplers per qubit, featuring the “Pegasus” connectivity graph between qubits. Since D-Wave can only produce binary solutions, we used the encoding as defined in Willsch et al. (2020) to convert the continuous variables *α*_*i*_ into *K* binary variables using base *B*:

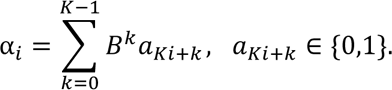

Using this encoding and also adding a penalty ξ to the loss function, the optimization problem becomes a quadratic unconstrained binary optimization (QUBO) problem, which can be run on a QA:

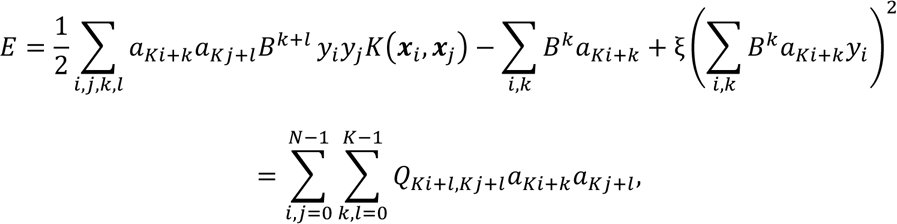

Where 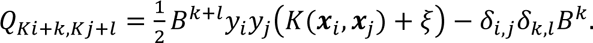. As the objective function above might necessitate connections between any pair of qubits, an embedding is necessary (Choi, 2008). Hyperparameters were selected using a custom 3-fold Monte-Carlo cross-validation on the training data. Hyperparameters included the type of kernel (linear versus Gaussian), *K* (between 2 and 10), ξ (between 2 and 6), ξ (between 0 and 5), and γ (between 2^−3^ and 2^3^). Closely related quantum ML approaches deploying QA have been used in classifying transcription factor binding to DNA (Li et al., 2018) and in classification of multiomics human cancer data (Li et al., 2021b).

#### Deep artificial neural network (DANN)

We adapted common deep learning methodologies to analyze genomic datasets (Alipanahi et al., 2015). Typical deep neural networks use a series of nonlinear transformations (termed layers), with the final output considered a prediction of class or regression variables. Each layer consists of a set of weights (*W*) and biases (*b*) that are tuned during the training phase to learn which nonlinear combinations of input features are most important for the prediction task. These types of models “automatically” learn patterns in the data and combine them in some abstract nonlinear fashion, to gain an ability to make predictions about the dataset.

The basic formulation of a fully connected DANN is given as

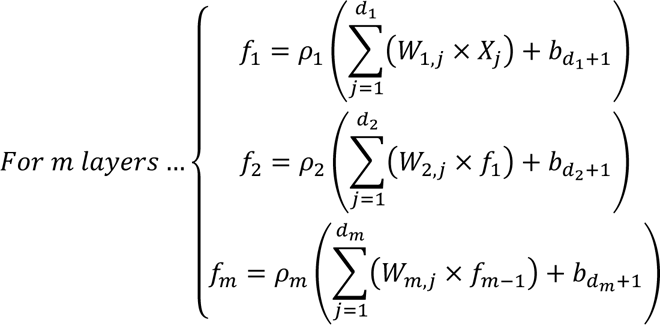

where the dimensions of *W* and *b* are determined by the number of neurons in each layer (*d*_1_, *d*_2_, …, *d*_*m*_). Each layer used rectified linear units as activation functions:

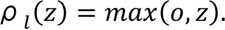

The final layer used a softmax function, with the number of neurons equal to the number of classes (ξ), to convert the logits to probabilities:

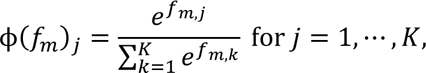

where *f*_*m,j*_ is the output of the *j*-th neuron of the *m*-th layer. In addition, we used the concept of “dropout,” which randomly sets a portion of input values (*η*) to the layer to zero during the training phase (Srivastava et al., 2014). This has a strong regularization effect (essentially by injecting random noise) that helps prevent models from overfitting. Layers that included dropout were formulated as

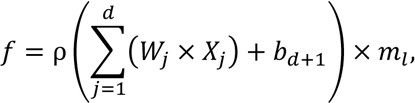

where *m*_*l*_ ∼ Bernoulli(η).

When evaluating models on test datasets, the dropout mask was not used. We used the categorical cross-entropy loss function to train DANNs, where (*K*_*n*_) is the minibatch size, *t*_*i*_ is the correct class index, and *p*_*i*_ is the class probability from the softmax layer:

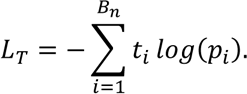

We used minibatch stochastic gradient descent with Nesterov momentum to update the DANN parameters based on the above-described loss function (Sutskever et al., 2013). We used the TensorFlow (Abadi et al., 2016) Python package to construct the DANNs.

#### Ensemble feature ranking

To derive an ensemble ranking of the feature importance, we first calculated the feature importances for each algorithm. LASSO, Ridge, SVM, and qSVM are linear models, and thus the feature importance was determined based on the value of the weight assigned to each feature, with a larger score corresponding to greater importance. RF creates a forest of decision trees, and as part of the fitting process, it determines an estimate of the feature importance by randomly permuting the features one at a time and determining the change in the accuracy. XGB calculates the feature importance by averaging the gain across all the trees, where the gain is the difference in the Gini purity of the parent node and the two children nodes.

The top 1000 most informative features of each model and for each partition of the data were retained. Because there were 100 partitions of the data, six algorithms (LASSO, Ridge, SVM, qSVM, RF, and XGB; DANN was not included because it lacks a robust approach to determine the feature importance), and up to 1000 features were retained, a total of up to 600000 possible features were considered for each feature set (“up to” since they might not be unique, as the top 1000 features for one partition of the data might exhibit some overlap with the top 1000 features for another partition of the data). We discarded the feature scores from an algorithm on any partition with a test AUROC < 0.7 in an attempt to exclude scores that might not truly be informative. To aggregate the scores, we scaled the scores by the most informative feature for each algorithm on each partition such that the feature scores were all between 0 and 1; i.e., for the first partition of the data, we scaled the 1000 most informative features from LASSO, then proceeded to do the same for Ridge, SVM, RF, and then repeated the process for each partition of the data. The scores were then averaged across all the partitions of the data to obtain a feature ranking for each method. If a feature was determined to be important for one partition of the data but not for others, it was given a value of 0 for all partitions of the data in which it did not appear. To determine a final ensemble feature ranking, the grand mean across all training partitions and algorithms was taken, and the features were sorted by the average score.

#### Structural causal modeling

We generated BBNs for the top 600 most informative genes as defined by ensemble feature ranking described above. BBNs were used to assess the conditional dependence and probabilistic relationships between the most informative genes. We used the TensorFlow (Abadi et al., 2016) Python package to construct the DANNs. We relied on a set of common assumptions to determine the causal structure: (1) causal sufficiency assumption, where there are no unobserved cofounders; (2) causal Markov assumption, where all d-separations in the graph (G) imply conditional independence in the observed probability distribution; and (3) causal faithfulness assumption, where all of the conditional independences in the observed probability distribution imply d-separations in the graph (*G*). We acknowledge that our data might not strictly meet all of these assumptions, however, the generated BBNs provide useful biological hypotheses that could be experimentally validated.

We determined BBNs using the bnlearn R package with the score-based hill-climbing algorithm that heuristically searched the optimality space of all possible DAGs (Scutari, 2010). As the hill-climbing algorithm can get trapped in local optima and is quite dependent on the starting structure, we initialized 100 BBNs starting from different network seeds. During the hill-climbing process, each candidate BBN was assessed with the Bayesian information criterion (BIC) score (Lam and Bacchus, 1994; Scutari, 2010): 

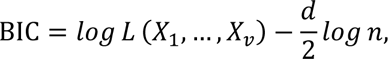

where *X*_1_, …, *X*_*v*_ is the node set, *d* is the number of free parameters, *n* is the sample size of the dataset, and *L* is the likelihood. Note that this definition of the BIC, which is the version implemented in the bnlearn package, rescales the classic definition by -2. The penalty term was used to prevent overly complicated structures and overfitting. Each run of the hill-climbing algorithm returns a structure that maximizes the BIC score (including evaluating the directions of edges). A caveat is that these structures might be partially oriented graphs (i.e., situations in which the directionality of some edges cannot be effectively determined). We use the *cextend* function from the *bnlearn* package to construct a DAG that is a consistent extension of *XX*. We then generated a consensus network based on the 100 networks after hill-climbing and selected to keep the edges that were present in graphs at least 30% of the time. Any residual undirected edges contained in the consensus network were discarded. We assessed the statistical significance of the edges within the imposed consensus network by randomly permuting the dataset 10000 times and evaluating the consensus structure on these scrambled datasets (thus providing an estimate of the null distribution). BBN edges with a FDR of at least 5% (i.e., the edge occurred in ≥500 of the random BBNs) were removed from the final network.

After deriving a final consensus network structure, we performed a series of *in silico* tests to determine the importance of each gene to the network. For each of the 600 genes, we removed all incident edges (both incoming and outgoing) and recalculated the BIC of the entire network. Doing so resulted in a lower BIC, and the magnitude of the change in BIC is a measure of how important a gene is to the network. We also experimented with permuting the data corresponding to a single gene; the results for the mean change in BIC using the permutation test and removing all the incident edges did not significantly differ (Pearson’s correlation > 0.999). Having derived a measure for the importance of each gene to the network, we can compare the mean change in BIC of the top five driver genes to 1000 random sets of five genes from the network.

### Real-time reverse transcription quantitative PCR (RT-qPCR)

Total RNA was extracted from cells using the RNeasy Mini Kit (Qiagen, Hilden, Germany), and the RNA quality was assessed using an Agilent 2100 BioAnalyzer before reverse transcription into cDNA with Maxima™ H Minus Mastermix and following the manufacturer’s instructions (Thermo Fisher Scientific, Waltham, MA, USA). RT-qPCR was performed using QuantStudio3 (Thermo Fisher Scientific, Waltham, MA, USA) according to the manufacturer’s protocol, and using PowerTrack™ SYBR™ Green Master Mix (Thermo Fisher Scientific, Waltham, MA, USA). The following primers were used: *ADAM9*, forward 5’-GGACTCAGAGGATTGCTGCATTTAG-3’, reverse 5’-CTTCGAAGTAGCTGAGTCATGCTGG-3’; and GAPDH (housekeeping gene), forward 5′-GGTGAAGGTCGGAGTCAACGGA-3′ and 5′-GAGGGATCTCGCTCCTGGAAGA-3′ (Integrated DNA Technologies, Coralville, IO, USA). The RT-qPCR protocol consisted of 95°C for 2 min followed by 40 cycles of 95°C for 5 s and 60°C for 30 s. All reactions were performed in duplicate, and the relative amounts of transcripts were calculated with the comparative Ct method. Gene expression changes were calculated using the 2^-ΔΔCt^ values calculated from averages of technical duplicates relative to the negative control. Melting-curve analysis was performed to assess the specificity of the PCR products.

### Enzyme-linked immunosorbent assays (ELISA)

The levels of soluble ADAM9 (sADAM9) and soluble MICA (sMICA) in the serum of critical and noncritical patients and healthy controls were quantified by ELISA. For soluble ADAM9, we used the Human sADAM9 DuoSet ELISA kit (R&D Systems, Minneapolis, MN, USA) following the manufacturer’s instructions. sMICA levels were measured with an in-house developed sandwich ELISA using two monoclonal mouse antibodies for capture (A13-C485B10 and A9-C255A9 at concentrations of 2 mg/ml and 0.2 mg/ml, respectively) and one biotinylated monoclonal mouse antibody for detection (A15-C199B9 at 60 pg/ml). Coating of MaxiSorp ELISA plates (Thermo Fisher Scientific, Waltham, MA, USA) was performed in PBS at 4°C overnight. After three washing steps with PBS, the wells were blocked with 200 μl of 10% BSA in PBS for 1 h at room temperature. All the following steps were carried out at room temperature with PBS/0.05% Tween 20/10% BSA, which was used as a diluent for all the reagents and sera. The plates were washed three times with PBS/0.05% Tween 20 between incubation steps. After blocking, the plates were incubated with 100 μl of sera, standards and controls for 2 h, followed by incubation with 100 μl of biotinylated detection antibody for 1 h. The plates were subsequently incubated for 1 h with 100 μl of a 5000-fold dilution of streptavidin poly-HRP (Thermo Fisher Scientific, Waltham, MA, USA) per well. The reactions were finally revealed using TMB Ultra (Thermo Fisher Scientific, Waltham, MA, USA) at 100 μl/well for 15 min and stopped with 100 μl of 1 M HCl. The absorbance was measured at 450 nm.

### Cell culture

Vero 76 cell lines were grown at 37 °C under 5% CO_2_ and maintained in DMEM (Thermo Fisher Scientific, Waltham, MA, USA) containing 100 units/ml penicillin and supplemented with 10% fetal bovine serum (Pan Biotech, Aidenbach, Germany). ACE2-expressing A549 cells (A549-ACE2) were grown at 37 °C under 5% CO_2_ and maintained in DMEM (Thermo Fisher Scientific, Waltham, MA, USA) containing 10 µg/ml of blasticidine S (Invitrogen, Carlsbad, CA, USA).

### Silencing and cell transfection

The cells were transfected with predesigned Stealth siRNA directed against ADAM9 (HSS112867) or the control Stealth RNAi Negative Control Duplex medium GC (45-55%) (Thermo Fisher Scientific, Waltham, MA, USA) using Lipofectamine^TM^ RNAiMAX Transfection Reagent (Thermo Fisher Scientific, Waltham, MA, USA). One day prior to transfection, the cells were seeded in a 24-well plate at 0.05 × 10^6^ cells per well. First, 1.5 μl of Lipofectamine^TM^ RNAiMAX Transfection Reagent was added to 25 μl of Opti-MEM^TM^ medium, followed by addition of the mix containing 5 pmoles of siRNA in 25 μl of Opti-MEM^TM^ medium (Thermo Fisher Scientific, Waltham, MA, USA). The mixture was incubated at room temperature for 5 min and then added to the cells. The cells were collected or infected after 48 h.

### Western blot

After collection and centrifugation, the cells were washed once in Dulbecco’s phosphate buffered saline (D-PBS, Sigma Aldrich, Saint-Louis, MI, USA). The pellet was resuspended in 60 μl of RIPA lysis buffer (150 mM NaCl, 5 mM EDTA, 1% NP40, 50 mM Tris pH 8, 0.5% sodium deoxycholate, and 0.1% SDS) including protease inhibitors (cOmplete, Roche Diagnostics, Rotkreuz, Switzerland) and maintained on ice for 20 min. The total cellular extract was then centrifuged for 30 min at 13000 g to remove all cell debris. A Bradford assay was used for protein quantification (Bio-Rad protein Assay, Bio-Rad Laboratories, Hercules, CA, USA). For Western blotting analysis, 20 μg of total cell extract was loaded on an 8% SDS-polyacrylamide gel. After migration, the proteins were transferred onto a PVDF membrane with a semidry transfer system (Trans-Blot, Bio-Rad Laboratories, Hercules, CA, USA). The membranes were blocked for 1 h in 5% skimmed milk/TBS 0.05%/Tween 20 and then incubated with the anti-ADAM9 antibody (ab218242; Abcam, Cambridge, UK) for 2 h at 4°C in 5% BSA/TBS 0.1% Tween at 1/1000 dilution. The membrane was then incubated with the secondary antibody coupled to HRP (Bio-Rad Laboratories, Hercules, CA, USA). Bound antibodies were revealed with an enhanced chemiluminescence detection system using ChemiDoc XRS (Bio-Rad Laboratories, Hercules, CA, USA). An anti-GAPDH antibody (MAB374, Merck Millipore, Burlington, MA, USA) was used for loading control.

### *In vitro* viral infections

Vero 76 and A549-ACE2 cell lines were infected with wild-type SARS-CoV-2 virus at Multiplicities Of Infections (MOIs) of 10 and 400, respectively. The percentage of infected cells was determined by staining with SARS-CoV-2 nucleocapsid (% of nucleocapsid positive cells), and virus released into the supernatant was analyzed by RT-PCR (copies/ml), after 2 and 3 days of infection for Vero 76 and A549-ACE2 cells, respectively.

### Flow cytometry staining

The cells were fixed for 20 min in 3.6% paraformaldehyde at 4°C, washed in 5% FCS in PBS and stained with anti-nucleocapsid antibody (GTX135357, Genetex, Irvine, CA, USA) at a 1/200 dilution in Perm/Wash^TM^ (Becton, Dickinson and Company, Franklin Lakes, NJ, USA) for 45 min at room temperature. The antibody was then revealed by incubation with an Alexa 647-labeled goat anti-rabbit monoclonal antibody (Ab150083, Abcam, Cambridge, UK) diluted 1/200 in 5% FCS in PBS for 45 min at room temperature.

### Viral RT-qPCR

RNA was extracted from the supernatant of infected cells using the NucleoSpin Dx Virus Kit (Macherey-Nagel GmbH & Co.KG, Düren, Germany). RT-qPCR was performed using TaqPath™ 1-Step RT-qPCR Master Mix (CG) on the Quanstudio3 instrument (Thermo Fisher Scientific, Waltham, MA, USA). The primer/probe mix used for absolute quantification of the virus was N1 and N2 from the 2019-nCoV RUO Kit (Integrated DNA Technologies, Coralville, IO, USA), and the positive control for the standard curve was 2019-nCoV N Positive Control (Integrated DNA Technologies, Coralville, IO, USA). The reaction was performed in 20 μl, which included 5 μl of eluted RNA, 5 μl of TaqPath Master Mix and 1.5 μl of the primer/probe. The RT-qPCR protocol consisted of 25°C for 2 min, 50°C for 15 min, and 95°C for 2 min, and 40 cycles of 95°C for 3 s and 60°C for 30 s. All reactions were performed in duplicate, and absolute quantification was calculated with the standard curve of the positive control.

## Data Availability

The data that support the findings of this study are available from the corresponding author upon reasonable request.

## Acknowledgments

We thank Drs. Benjamin Terrier, and Philippe Georgel for critical reading of this manuscript, Olivier Schwartz and his team for sharing the A549*-*ACE2 cell line and Christel Thauvin-Robinet and Franck Lethimonnier (AVIESAN) for providing access to the AURAGEN PFMG2025 platform. We would like to acknowledge the Cytometry Core Facility Hyperion (Brest, France) for their technical assistance as well as colleagues from the Immunophenotyping Module of CIPHE (Centre d’Immunophénomique - CIPHE (PHENOMIN), Aix Marseille Université (UMS3367), Inserm (US012), CNRS (UMS3367), Marseille, France) for acquiring the mass cytometry dataset used in this work. We thank Sweta R. Bajaj, Chen-Hao Chen, and Allison R. McLean for the bioinformatics processing of patient data, Emmanuelle Jouanguy for help with anti-IFN autoantibodies assessments, Cecilia Hellström for expert technical and bioinformatics assistance and Léa Resmini for technical assistance.

## Author Contributions

Conceptualization: RC, TWC, SB, ; Methodology: RC, RL, CC, SG, RG, RM, DL, CMo, TWC, SB, ; Software: RL, SG, RG, JML, RM, HW, NP, SM, TS, AV, DL, TWC, ; Validation: RC, RL, CC, SG, RG, RM, HW, TWC, SB, ; Formal analysis: RC, RL, CC, SG, RG, JML, RM, PB, HW, NP, SM, TS, SK, MJA, EP, AV, PLT, DL, JRG, CMo, TWC, SB, ; Investigation: RC, CC, VR, PS, AH, LM, PB, AG, QZ, AM, AP, AB, OT, SBT, AL, GL, JL, SS, Aha, CM, SK, MJA, EP, PN, AT, PLT, MR, CMo, SB, ; Resources: RC, JH, CC, RM, FD, YR, MSD, OC, TNCT, XM, OH, SFK, BG, BD, KK, JP, PMM, SK, MJA, EP, PN, AT, DS, FS, PLT, JLC, YH, DL, JRG, FM, CMo, TWC, SB, ; Data Curation: RC, RL, CC, SG, RG, JML, RM, TWC, SB, ; Writing - Original Draft: RC, RL, TWC, SB, ; Writing - Review & Editing: RC, RL, JH, CC, VR, RG, PS, RM, AH, LM, FD, YR, TNCT, AP, MS, JP, SK, MJA, EP, JS, JLC, DL, MR, JRG, CMo, TWC, SB, ; Visualization: RC, RL, RG, TWC, ; Supervision: RC, JRG, CMo, TWC, SB, ; Project administration: RC, AM, MR, CMo, TWC, SB, ; Funding acquisition: RC, JRG, TWC, SB. All authors have read and agreed to the content of the manuscript.

## REFERENCES

1. Abadi, M., Barham, P., Chen, J., Chen, Z., Davis, A., Dean, J., Devin, M., Ghemawat, S., Irving, G., and Isard, M. (2016). Tensorflow: A system for large-scale machine learning. In 12th {USENIX} Symposium on Operating Systems Design and Implementation ({OSDI} 16), pp. 265–283.

2. Al-Samkari, H., Karp Leaf, R.S., Dzik, W.H., Carlson, J.C.T., Fogerty, A.E., Waheed, A., Goodarzi, K., Bendapudi, P.K., Bornikova, L., Gupta, S., et al. (2020). COVID-19 and coagulation: Bleeding and thrombotic manifestations of SARS-CoV-2 infection. Blood 136, 489–500.

3. Alipanahi, B., Delong, A., Weirauch, M.T., and Frey, B.J. (2015). Predicting the sequence specificities of DNA-and RNA-binding proteins by deep learning. Nat. Biotechnol. 33, 831– 838.

4. Arunachalam, P.S., Wimmers, F., Mok, C.K.P., Perera, R.A.P.M., Scott, M., Hagan, T., Sigal, N., Feng, Y., Bristow, L., Tsang, O.T.Y., et al. (2020). Systems biological assessment of immunity to mild versus severe COVID-19 infection in humans. Science. 369, 1210–1220.

5. Van der Auwera, G.A., Carneiro, M.O., Hartl, C., Poplin, R., Del Angel, G., Levy-Moonshine, A., Jordan, T., Shakir, K., Roazen, D., and Thibault, J. (2013). From FastQ data to high-confidence variant calls: the genome analysis toolkit best practices pipeline. Curr. Protoc. Bioinforma. 43, 11.10. 1–11.10. 33.

6. Bastard, P., Rosen, L.B., Zhang, Q., Michailidis, E., Hoffmann, H.H., Zhang, Y., Dorgham, K., Philippot, Q., Rosain, J., Béziat, V., et al. (2020). Autoantibodies against type I IFNs in patients with life-threatening COVID-19. Science. 370, eabd4585.

7. Bastard, P., Orlova, E., Sozaeva, L., Lévy, R., James, A., Schmitt, M.M., Ochoa, S., Kareva, M., Rodina, Y., Gervais, A., et al. (2021). Preexisting autoantibodies to type I IFNs underlie critical COVID-19 pneumonia in patients with APS-1. J. Exp. Med. 218, 29.

8. Bazzone, L.E., King, M., MacKay, C.R., Kyawe, P.P., Meraner, P., Lindstrom, D., Rojas-Quintero, J., Owen, C.A., Wang, J.P., Brass, A.L., et al. (2019). A disintegrin and metalloproteinase 9 domain (ADAM9) is a major susceptibility factor in the early stages of encephalomyocarditis virus infection. MBio 10, e02734–18.

9. De Biasi, S., Lo Tartaro, D., Meschiari, M., Gibellini, L., Bellinazzi, C., Borella, R., Fidanza, L., Mattioli, M., Paolini, A., Gozzi, L., et al. (2020a). Expansion of plasmablasts and loss of memory B cells in peripheral blood from COVID-19 patients with pneumonia. Eur. J. Immunol. 50, 1283–1294.

10. De Biasi, S., Meschiari, M., Gibellini, L., Bellinazzi, C., Borella, R., Fidanza, L., Gozzi, L., Iannone, A., Lo Tartaro, D., Mattioli, M., et al. (2020b). Marked T cell activation, senescence, exhaustion and skewing towards TH17 in patients with COVID-19 pneumonia. Nat. Commun. 11, 1–17.

11. Boser, B.E., Guyon, I.M., and Vapnik, V.N. (1992). A training algorithm for optimal margin classifiers. In Proceedings of the Fifth Annual Workshop on Computational Learning Theory, (ACM), pp. 144–152.

12. Breiman, L. (2001). Random forests. Mach. Learn. 45, 5–32.

13. Breiman, L., Friedman, J., Stone, C.J., and Olshen, R.A. (1993). Classification and regression trees (Chapman & Hall). 11, 246–280.

14. Buchrieser, J., Dufloo, J., Hubert, M., Monel, B., Planas, D., Rajah, M.M., Planchais, C., Porrot, F., Guivel-Benhassine, F., Van der Werf, S., et al. (2020). Syncytia formation by SARS-CoV-2-infected cells. EMBO J. 39, e106267.

15. Carapito, R., and Bahram, S. (2015). Genetics, genomics, and evolutionary biology of NKG2D ligands. Immunol. Rev. 267, 88–116.

16. CDC (2021). Scientific Evidence for Conditions that Increase Risk of Severe Illness | COVID-19 | CDC. Cent. Dis. Control Prev.

17. Chen, T., and Guestrin, C. (2016). Xgboost: A scalable tree boosting system. In Proceedings of the 22nd Acm Sigkdd International Conference on Knowledge Discovery and Data Mining, pp. 785–794.

18. Chen, G., Wu, D., Guo, W., Cao, Y., Huang, D., Wang, H., Wang, T., Zhang, X., Chen, H., Yu, H., et al. (2020a). Clinical and immunological features of severe and moderate coronavirus disease 2019. J. Clin. Invest. 130, 2620–2629.

19. Chen, L., Long, X., Xu, Q., Tan, J., Wang, G., Cao, Y., Wei, J., Luo, H., Zhu, H., Huang, L., et al. (2020b). Elevated serum levels of S100A8/A9 and HMGB1 at hospital admission are correlated with inferior clinical outcomes in COVID-19 patients. Cell. Mol. Immunol. 17, 992– 994.

20. Chen, N., Zhou, M., Dong, X., Qu, J., Gong, F., Han, Y., Qiu, Y., Wang, J., Liu, Y., Wei, Y., et al. (2020c). Epidemiological and clinical characteristics of 99 cases of 2019 novel coronavirus pneumonia in Wuhan, China: a descriptive study. Lancet 395, 507–513.

21. Choi, V. (2008). Minor-embedding in adiabatic quantum computation: I. The parameter setting problem. Quantum Inf. Process. 7, 193–209.

22. Chou, C.-W., Huang, Y.-K., Kuo, T.-T., Liu, J.-P., and Sher, Y.-P. (2020). An Overview of ADAM9: Structure, Activation, and Regulation in Human Diseases. Int. J. Mol. Sci. 21, 7790.

23. Chua, R.L., Lukassen, S., Trump, S., Hennig, B.P., Wendisch, D., Pott, F., Debnath, O., Thürmann, L., Kurth, F., Völker, M.T., et al. (2020). COVID-19 severity correlates with airway epithelium–immune cell interactions identified by single-cell analysis. Nat. Biotechnol. 38, 970–979.

24. Cortes, and Vapnik V. (1995). Support-vector networks. Mach. Learn. 20, 273–297.

25. Davies, N.G., Jarvis, C.I., van Zandvoort, K., Clifford, S., Sun, F.Y., Funk, S., Medley, G., Jafari, Y., Meakin, S.R., Lowe, R., et al. (2021). Increased mortality in community-tested cases of SARS-CoV-2 lineage B.1.1.7. Nature. in press.

26. Dejnirattisai, W., Zhou, D., Supasa, P., Liu, C., Mentzer, A.J., Ginn, H.M., Zhao, Y., Duyvesteyn, H.M.E., Tuekprakhon, A., Nutalai, R., et al. (2021). Antibody evasion by the P.1 strain of SARS-CoV-2. Cell. in press.

27. DePristo, M.A., Banks, E., Poplin, R., Garimella, K. V, Maguire, J.R., Hartl, C., Philippakis, A.A., Del Angel, G., Rivas, M.A., and Hanna, M. (2011). A framework for variation discovery and genotyping using next-generation DNA sequencing data. Nat. Genet. 43, 491–498.

28. Deutsch, E.W., Csordas, A., Sun, Z., Jarnuczak, A., Perez-Riverol, Y., Ternent, T., Campbell, D.S., Bernal-Llinares, M., Okuda, S., Kawano, S., et al. (2017). The ProteomeXchange consortium in 2017: Supporting the cultural change in proteomics public data deposition. Nucleic Acids Res. 45, D1100–D1106.

29. Dobin, A., Davis, C.A., Schlesinger, F., Drenkow, J., Zaleski, C., Jha, S., Batut, P., Chaisson, M., and Gingeras, T.R. (2013). STAR: ultrafast universal RNA-seq aligner. Bioinformatics 29, 15–21.

30. Giamarellos-Bourboulis, E.J., Netea, M.G., Rovina, N., Akinosoglou, K., Antoniadou, A., Antonakos, N., Damoraki, G., Gkavogianni, T., Adami, M.E., Katsaounou, P., et al. (2020). Complex Immune Dysregulation in COVID-19 Patients with Severe Respiratory Failure. Cell Host Microbe 27, 992–1000.e3.

31. Glebov, O.O. (2020). Understanding SARS-CoV-2 endocytosis for COVID-19 drug repurposing. FEBS J. 287, 3664–3671.

32. Gordon, D.E., Jang, G.M., Bouhaddou, M., Xu, J., Obernier, K., White, K.M., O’Meara, M.J., Rezelj, V. V., Guo, J.Z., Swaney, D.L., et al. (2020a). A SARS-CoV-2 protein interaction map reveals targets for drug repurposing. Nature 583, 459–468.

33. Gordon, D.E., Hiatt, J., Bouhaddou, M., Rezelj, V. V., Ulferts, S., Braberg, H., Jureka, A.S., Obernier, K., Guo, J.Z., Batra, J., et al. (2020b). Comparative host-coronavirus protein interaction networks reveal pan-viral disease mechanisms. Science. 370, eabe9403.

34. Guan, W., Ni, Z., Hu, Y., Liang, W., Ou, C., He, J., Liu, L., Shan, H., Lei, C., Hui, D.S.C., et al. (2020). Clinical Characteristics of Coronavirus Disease 2019 in China. N. Engl. J. Med. 382, 1708–1720.

35. Hadjadj, J., Yatim, N., Barnabei, L., Corneau, A., Boussier, J., Smith, N., Péré, H., Charbit, B., Bondet, V., Chenevier-Gobeaux, C., et al. (2020). Impaired type I interferon activity and inflammatory responses in severe COVID-19 patients. Science. 369, 718–724.

36. Helms, J., Tacquard, C., Severac, F., Leonard-Lorant, I., Ohana, M., Delabranche, X., Merdji, H., Clere-Jehl, R., Schenck, M., Fagot Gandet, F., et al. (2020). High risk of thrombosis in patients with severe SARS-CoV-2 infection: a multicenter prospective cohort study. Intensive Care Med. 46, 1089–1098.

37. Hermine, O., Mariette, X., Tharaux, P.L., Resche-Rigon, M., Porcher, R., and Ravaud, P. (2021). Effect of Tocilizumab vs Usual Care in Adults Hospitalized with COVID-19 and Moderate or Severe Pneumonia: A Randomized Clinical Trial. JAMA Intern. Med. 181, 32– 40.

38. Hicks, S., Loo, D., Sinkevicius, K., Scribner, J., Barat, B., Yoder, N., Espelin, C., Themeles, M., Chen, F., Lucas, J., et al. (2019). Abstract 1533: IMGC936, a first-in-class ADAM9-targeting antibody-drug conjugate, demonstrates promising anti-tumor activity. In Cancer Research, (American Association for Cancer Research (AACR)), pp. 1533–1533.

39. Hoerl, A.E., and Kennard, R.W. (1970). Ridge regression: Biased estimation for nonorthogonal problems. Technometrics 12, 55–67.

40. Hoerl, A.E., Kannard, R.W., and Baldwin, K.F. (1975). Ridge regression: some simulations. Commun. Stat. Methods 4, 105–123.

41. Horton, R. (2020). Offline: COVID-19 is not a pandemic. Lancet 396, 874.

42. Benchmarking - COVID-19 - UTIs Brasileiras. http://Www.Utisbrasileiras.Com.Br/En/Covid-19/Benchmarking-Covid-19/. Accessed 9/5/2021.

43. Huang, C., Wang, Y., Li, X., Ren, L., Zhao, J., Hu, Y., Zhang, L., Fan, G., Xu, J., Gu, X., et al. (2020). Clinical features of patients infected with 2019 novel coronavirus in Wuhan, China. Lancet 395, 497–506.

44. Huber, W., von Heydebreck, A., Sultmann, H., Poustka, A., and Vingron, M. (2002). Variance stabilization applied to microarray data calibration and to the quantification of differential expression. Bioinformatics 18, S96–S104.

45. Institut Pasteur (2021). Protocol: Real-time RT-PCR assays for the detection of SARS-CoV-2. https://www.who.int/docs/default-source/coronaviruse/real-time-rt-pcr-assays-for-thedetection-of-sars-cov-2-institut-pasteur-paris.pdf?sfvrsn=3662fcb6_2. Accessed 9/5/2021.

46. Ioannidis, J.P.A., Axfors, C., and Contopoulos-Ioannidis, D.G. (2020). Population-level COVID-19 mortality risk for non-elderly individuals overall and for non-elderly individuals without underlying diseases in pandemic epicenters. Environ. Res. 188, 109890.

47. Johnson, M.W., Amin, M.H.S., Gildert, S., Lanting, T., Hamze, F., Dickson, N., Harris, R., Berkley, A.J., Johansson, J., and Bunyk, P. (2011). Quantum annealing with manufactured spins. Nature 473, 194–198.

48. Kendig, K.I., Baheti, S., Bockol, M.A., Drucker, T.M., Hart, S.N., Heldenbrand, J.R., Hernaez, M., Hudson, M.E., Kalmbach, M.T., and Klee, E.W. (2019). Sentieon DNASeq variant calling workflow demonstrates strong computational performance and accuracy. Front. Genet. 10, 1–7.

49. Klok, F.A., Kruip, M.J.H.A., van der Meer, N.J.M., Arbous, M.S., Gommers, D.A.M.P.J., Kant, K.M., Kaptein, F.H.J., van Paassen, J., Stals, M.A.M., Huisman, M. V., et al. (2020a). Incidence of thrombotic complications in critically ill ICU patients with COVID-19. Thromb. Res. 191, 145–147.

50. Klok, F.A., Kruip, M.J.H.A., van der Meer, N.J.M., Arbous, M.S., Gommers, D., Kant, K.M., Kaptein, F.H.J., van Paassen, J., Stals, M.A.M., Huisman, M. V., et al. (2020b). Confirmation of the high cumulative incidence of thrombotic complications in critically ill ICU patients with COVID-19: An updated analysis. Thromb. Res. 191, 148–150.

51. Kohga, K., Takehara, T., Tatsumi, T., Ishida, H., Miyagi, T., Hosui, A., and Hayashi, N. (2010). Sorafenib inhibits the shedding of major histocompatibility complex class i-related chain a on hepatocellular carcinoma cells by down-regulating a disintegrin and metalloproteinase 9. Hepatology 51, 1264–1273.

52. Kokla, M., Virtanen, J., Kolehmainen, M., Paananen, J., and Hanhineva, K. (2019). Random forest-based imputation outperforms other methods for imputing LC-MS metabolomics data: A comparative study. BMC Bioinformatics 20, 492.

53. Kuhn, M. (2008). Building predictive models in R using the caret package. J. Stat. Softw. 28, 1–26.

54. Lam, W., and Bacchus, F. (1994). Learning Bayesian belief networks: An approach based on the MDL principle. Comput. Intell. 10, 269–293.

55. Leisman, D.E., Ronner, L., Pinotti, R., Taylor, M.D., Sinha, P., Calfee, C.S., Hirayama, A. V., Mastroiani, F., Turtle, C.J., Harhay, M.O., et al. (2020). Cytokine elevation in severe and critical COVID-19: a rapid systematic review, meta-analysis, and comparison with other inflammatory syndromes. Lancet Respir. Med. 8, 1233–1244.

56. Li, B., and Dewey, C.N. (2011). RSEM: accurate transcript quantification from RNA-Seq data with or without a reference genome. BMC Bioinformatics 12, 323.

57. Li, H., and Durbin, R. (2009). Fast and accurate short read alignment with Burrows–Wheeler transform. Bioinformatics 25, 1754–1760.

58. Li, J., Guo, M., Tian, X., Wang, X., Yang, X., Wu, P., Liu, C., Xiao, Z., Qu, Y., Yin, Y., et al. (2021a). Virus-Host Interactome and Proteomic Survey Reveal Potential Virulence Factors Influencing SARS-CoV-2 Pathogenesis. Med 2, 99–112.e7.

59. Li, K., Wang, S.W., Li, Y., Martin, R.E., Li, L., Lu, M., Lamhamedi-Cherradi, S.E., Hu, G., Demissie-Sanders, S., Zheng, J., et al. (2005). Identification and expression of a new type II transmembrane protein in human mast cells. Genomics 86, 68–75.

60. Li, R.Y., Di Felice, R., Rohs, R., and Lidar, D.A. (2018). Quantum annealing versus classical machine learning applied to a simplified computational biology problem. Npj Quantum Inf. 4. 14.

61. Li, R.Y., Gujja, S., Bajaj, S.R., Gamel, O.E., Cilfone, N., Gulcher, J.R., Lidar, D.A., and Chittenden, T.W. (2021b). Quantum processor-inspired machine learning in the biomedical sciences. Patterns 0, 100246.

62. Li, X., Marmar, T., Xu, Q., Tu, J., Yin, Y., Tao, Q., Chen, H., Shen, T., and Xu, D. (2020). Predictive indicators of severe COVID-19 independent of comorbidities and advanced age: A nested case -control study. Epidemiol. Infect. 148.

63. Liu, Z., Xu, E., Zhao, H.T., Cole, T., and West, A.B. (2020). LRRK2 and Rab10 coordinate macropinocytosis to mediate immunological responses in phagocytes. EMBO J. 39.

64. Lucas, C., Wong, P., Klein, J., Castro, T.B.R., Silva, J., Sundaram, M., Ellingson, M.K., Mao, T., Oh, J.E., Israelow, B., et al. (2020). Longitudinal analyses reveal immunological misfiring in severe COVID-19. Nature 584, 463–469.

65. Van Der Made, C.I., Simons, A., Schuurs-Hoeijmakers, J., Van Den Heuvel, G., Mantere, T., Kersten, S., Van Deuren, R.C., Steehouwer, M., Van Reijmersdal, S. V., Jaeger, M., et al. (2020). Presence of Genetic Variants among Young Men with Severe COVID-19. JAMA - J. Am. Med. Assoc. 324, 663–673.

66. Mariette, X., Hermine, O., Resche-Rigon, M., Porcher, R., Ravaud, P., Bureau, S., Dougados, M., Tibi, A., Azoulay, E., Cadranel, J., et al. (2021). Effect of anakinra versus usual care in adults in hospital with COVID-19 and mild-to-moderate pneumonia (CORIMUNO-ANA-1): a randomised controlled trial. Lancet Respir. Med. 9, 295–304.

67. Mehta, P., McAuley, D.F., Brown, M., Sanchez, E., Tattersall, R.S., and Manson, J.J. (2020). COVID-19: consider cytokine storm syndromes and immunosuppression. Lancet 395, 1033– 1034.

68. Meizlish, M.L., Pine, A.B., Bishai, J.D., Goshua, G., Nadelmann, E.R., Simonov, M., Chang, C.-H., Zhang, H., Shallow, M., Bahel, P., et al. (2021). A neutrophil activation signature predicts critical illness and mortality in COVID-19. Blood Adv. 5, 1164–1177.

69. Messner, C.B., Demichev, V., Wendisch, D., Michalick, L., White, M., Freiwald, A., Textoris-Taube, K., Vernardis, S.I., Egger, A.S., Kreidl, M., et al. (2020). Ultra-High-Throughput Clinical Proteomics Reveals Classifiers of COVID-19 Infection. Cell Syst. 11, 11–24.e4.

70. Odak, I., Barros-Martins, J., Bošnjak, B., Stahl, K., David, S., Wiesner, O., Busch, M., Hoeper, M.M., Pink, I., Welte, T., et al. (2020). Reappearance of effector T cells is associated with recovery from COVID-19. EBioMedicine 57, 102885.

71. Quah, P., Li, A., Phua, J., and Phua, J. (2020). Mortality rates of patients with COVID-19 in the intensive care unit: A systematic review of the emerging literature. Crit. Care 24, 285.

72. Ranieri, V.M., Rubenfeld, G.D., Thompson, B.T., Ferguson, N.D., Caldwell, E., Fan, E., Camporota, L., and Slutsky, A.S. (2012). Acute respiratory distress syndrome: The Berlin definition. JAMA - J. Am. Med. Assoc. 307, 2526–2533.

73. Remy, K.E., Mazer, M., Striker, D.A., Ellebedy, A.H., Walton, A.H., Unsinger, J., Blood, T.M., Mudd, P.A., Yi, D.J., Mannion, D.A., et al. (2020). Severe immunosuppression and not a cytokine storm characterizes COVID-19 infections. JCI Insight 5, e140329.

74. Ricard, N., Scott, R.P., Booth, C.J., Velazquez, H., Cilfone, N.A., Baylon, J.L., Gulcher, J.R., Quaggin, S.E., Chittenden, T.W., and Simons, M. (2019). Endothelial ERK1/2 signaling maintains integrity of the quiescent endothelium. J. Exp. Med. 216, 1874–1890.

75. Ritchie, M.E., Phipson, B., Wu, D., Hu, Y., Law, C.W., Shi, W., and Smyth, G.K. (2015). Limma powers differential expression analyses for RNA-sequencing and microarray studies. Nucleic Acids Res. 43, e47.

76. Robinson, M.D., and Oshlack, A. (2010). A scaling normalization method for differential expression analysis of RNA-seq data. Genome Biol. 11, R25.

77. Robinson, M.D., McCarthy, D.J., and Smyth, G.K. (2010). edgeR: a Bioconductor package for differential expression analysis of digital gene expression data. Bioinformatics 26, 139– 140.

78. Rubin, E.J., Longo, D.L., and Baden, L.R. (2021). Interleukin-6 Receptor Inhibition in Covid-19 — Cooling the Inflammatory Soup. N. Engl. J. Med. 384, 1564–1565.

79. Rudberg, A.S., Havervall, S., Månberg, A., Jernbom Falk, A., Aguilera, K., Ng, H., Gabrielsson, L., Salomonsson, A.C., Hanke, L., Murrell, B., et al. (2020). SARS-CoV-2 exposure, symptoms and seroprevalence in healthcare workers in Sweden. Nat. Commun. 11, 1–8.

80. Sánchez-Cerrillo, I., Landete, P., Aldave, B., Sánchez-Alonso, S., Sánchez-Azofra, A., Marcos-Jiménez, A., Ávalos, E., Alcaraz-Serna, A., de los Santos, I., Mateu-Albero, T., et al. (2020). COVID-19 severity associates with pulmonary redistribution of CD1c+ DCs and inflammatory transitional and nonclassical monocytes. J. Clin. Invest. 130, 6290–6300.

81. Sanyal, R., Polyak, M.J., Zuccolo, J., Puri, M., Deng, L., Roberts, L., Zuba, A., Storek, J., Luider, J.M., Sundberg, E.M., et al. (2017). MS4A4A: a novel cell surface marker for M2 macrophages and plasma cells. Immunol. Cell Biol. 95, 611–619.

82. Schulte-Schrepping, J., Reusch, N., Paclik, D., Baßler, K., Schlickeiser, S., Zhang, B., Krämer, B., Krammer, T., Brumhard, S., Bonaguro, L., et al. (2020). Severe COVID-19 Is Marked by a Dysregulated Myeloid Cell Compartment. Cell 182, 1419–1440.e23.

83. Scutari, M. (2010). Learning Bayesian Networks with the bnlearn R Package. 2010 *35*, 22.

84. Shabalin, A.A. (2012). Matrix eQTL: ultra fast eQTL analysis via large matrix operations. Bioinformatics 28, 1353–1358.

85. Shelton, J.F., Shastri, A.J., Ye, C., Weldon, C.H., Filshtein-Sonmez, T., Coker, D., Symons, A., Esparza-Gordillo, J., 23andMe COVID-19 Team, Aslibekyan, S., et al. (2021). Trans-ancestry analysis reveals genetic and nongenetic associations with COVID-19 susceptibility and severity. Nat. Genet. in press.

86. Shen, B., Yi, X., Liu, H., Chen, H., and Guo Correspondence, T. (2020). Proteomic and Metabolomic Characterization of COVID-19 Patient Sera. Cell 182, 59–72.e15.

87. Shu, T., Ning, W., Wu, D., Xu, J., Han, Q., Huang, M., Zou, X., Yang, Q., Yuan, Y., Bie, Y., et al. (2020). Plasma Proteomics Identify Biomarkers and Pathogenesis of COVID-19. Immunity 53, 1108–1122.e5.

88. Silvin, A., Chapuis, N., Dunsmore, G., Goubet, A.G., Dubuisson, A., Derosa, L., Almire, C., Hénon, C., Kosmider, O., Droin, N., et al. (2020). Elevated Calprotectin and Abnormal Myeloid Cell Subsets Discriminate Severe from Mild COVID-19. Cell 182, 1401–1418.e18.

89. Srivastava, N., Hinton, G., Krizhevsky, A., Sutskever, I., and Salakhutdinov, R. (2014). Dropout: a simple way to prevent neural networks from overfitting. J. Mach. Learn. Res. 15, 1929–1958.

90. Su, Y., Chen, D., Yuan, D., Lausted, C., Choi, J., Dai, C.L., Voillet, V., Duvvuri, V.R., Scherler, K., Troisch, P., et al. (2020). Multi-Omics Resolves a Sharp Disease-State Shift between Mild and Moderate COVID-19. Cell 183, 1479–1495.e20.

91. Sui, Y., and Zeng, W. (2020). MS4A4A Regulates Arginase 1 Induction during Macrophage Polarization and Lung Inflammation in Mice. Eur. J. Immunol. 50, 1602–1605.

92. Sutskever, I., Martens, J., Dahl, G., and Hinton, G. (2013). On the importance of initialization and momentum in deep learning. In International Conference on Machine Learning, pp. 1139–1147.

93. The Severe Covid-19 GWAS (2020). Genomewide Association Study of Severe Covid-19 with Respiratory Failure. N. Engl. J. Med. 383, 1522–1534.

94. Tibshirani, R. (1996). Regression shrinkage and selection via the lasso. J. R. Stat. Soc. Ser. B 58, 267–288.

95. Trouillet-Assant, S., Viel, S., Gaymard, A., Pons, S., Richard, J.C., Perret, M., Villard, M., Brengel-Pesce, K., Lina, B., Mezidi, M., et al. (2020). Type I IFN immunoprofiling in COVID-19 patients. J. Allergy Clin. Immunol. 146, 206–208.e2.

96. Wang, P., Sha, J., Meng, M., Wang, C., Yao, Q., Zhang, Z., Sun, W., Wang, X., Qie, G., Bai, X., et al. (2020). Risk factors for severe COVID-19 in middle-aged patients without comorbidities: a multicentre retrospective study. J. Transl. Med. 18, 461.

97. Wei, L., Wang, W., Chen, D., and Xu, B. (2020). Dysregulation of the immune response affects the outcome of critical COVID-19 patients. J. Med. Virol. 92, 2768–2776.

98. Willsch, D., Willsch, M., De Raedt, H., and Michielsen, K. (2020). Support vector machines on the D-Wave quantum annealer. Comput. Phys. Commun. 248, 107006.

99. Xie, W., Chen, L., Chen, L., and Kou, Q. (2020). Silencing of long non-coding RNA MALAT1 suppresses inflammation in septic mice: role of microRNA-23a in the down-regulation of MCEMP1 expression. Inflamm. Res. 69, 179–190.

100. Yu, P., Wilhelm, K., Dubrac, A., Tung, J.K., Alves, T.C., Fang, J.S., Xie, Y., Zhu, J., Chen, Z., De Smet, F., et al. (2017). FGF-dependent metabolic control of vascular development. Nature 545, 224–241.

101. Zhang, Q., Liu, Z., Moncada-Velez, M., Chen, J., Ogishi, M., Bigio, B., Yang, R., Arias, A.A., Zhou, Q., Han, J.E., et al. (2020). Inborn errors of type I IFN immunity in patients with life-threatening COVID-19. Science. 370, eabd4570.

102. Zhou, Z., Ren, L., Zhang, L., Zhong, J., Xiao, Y., Jia, Z., Guo, L., Yang, J., Wang, C., Jiang, S., et al. (2020). Overly Exuberant Innate Immune Response to SARS-CoV-2 Infection. SSRN Electron. J.

