## supplemental file for "Identification of driver genes for severe forms of COVID-19 in a deeply phenotyped young patient cohort"

#### SUPPLEMENTAL INFORMATION TITLES AND LEGENDS

##### Table S1. Critical patients with detectable viremia and the corresponding FPKM values of SARS-CoV-2 gene

| Sample ID | FPKM* mean | ORF1ab | ORF1ab | S | N | ORF3a | M | ORF8 | ORF7a | E | ORF6 | ORF7b | ORF10 |
|---|---|---|---|---|---|---|---|---|---|---|---|---|---|
| P14 | 0.0008333 | 0 | 0.01 | 0 | 0 | 0 | 0 | 0 | 0 | 0 | 0 | 0 | 0 |
| P27 | 0.0008333 | 0 | 0.01 | 0 | 0 | 0 | 0 | 0 | 0 | 0 | 0 | 0 | 0 |
| P31 | 0.0125 | 0 | 0 | 0.01 | 0 | 0 | 0 | 0 | 0.14 | 0 | 0 | 0 | 0 |
| P32 | 0.0025 | 0 | 0 | 0 | 0.03 | 0 | 0 | 0 | 0 | 0 | 0 | 0 | 0 |
| P37 | 0.2683333 | 0.14 | 0 | 0.18 | 0.41 | 0.08 | 0.52 | 0.13 | 0.13 | 0.35 | 1.28 | 0 | 0 |
| P39 | 0.0175 | 0 | 0.01 | 0.03 | 0 | 0 | 0.05 | 0 | 0.12 | 0 | 0 | 0 | 0 |
| P43 | 0.0066667 | 0.01 | 0 | 0 | 0.07 | 0 | 0 | 0 | 0 | 0 | 0 | 0 | 0 |
| P46 | 0.02 | 0.02 | 0 | 0.04 | 0.15 | 0.03 | 0 | 0 | 0 | 0 | 0 | 0 | 0 |

*FPKM: fragments per kilo per million

**Table S2. Performance metrics on the training and testing sets of each algorithm in the ensemble computational intelligence approach**

| | LASSO | Ridge | SVM | qSVM | XGB | RF | DANN |
|---|---|---|---|---|---|---|---|
| **Accuracy (Train/Test)** | 0.9991 ± 0.0004 / 0.9677 ± 0.0050 | 1.0000 ± 0.0000 / 0.9169 ± 0.0072 | 1.0000 ± 0.0000 / 0.9223 ± 0.0075 | 0.9245 ± 0.0028 / 0.8677 ± 0.0121 | 0.9952 ± 0.0008 / 0.9146 ± 0.0076 | 1.0000 ± 0.0000 / 0.9254 ± 0.0072 | 1.0000 ± 0.0000 / 0.9131 ± 0.0083 |
| **Balanced Acc. (Train/Test)** | 0.9987 ± 0.0006 / 0.9503 ± 0.0078 | 1.0000 ± 0.0000 / 0.8990 ± 0.0094 | 1.0000 ± 0.0000 / 0.9068 ± 0.0092 | 0.9189 ± 0.0039 / 0.8607 ± 0.0118 | 0.9930 ± 0.0012 / 0.8932 ± 0.0100 | 1.0000 ± 0.0000 / 0.9072 ± 0.0094 | 1.0000 ± 0.0000 / 0.9032 ± 0.0097 |
| **AUROC (Train/Test)** | 1.0000 ± 0.0000 / 0.9908 ± 0.0036 | 1.0000 ± 0.0000 / 0.9547 ± 0.0075 | 1.0000 ± 0.0000 / 0.9633 ± 0.0070 | 0.9667 ± 0.0029 / 0.9386 ± 0.0081 | 0.9999 ± 0.0000 / 0.9443 ± 0.0079 | 1.0000 ± 0.0000 / 0.9360 ± 0.0091 | 1.0000 ± 0.0000 / 0.9435 ± 0.0081 |
| **F1 (Train/Test)** | 0.9993 ± 0.0003 / 0.9780 ± 0.0034 | 1.0000 ± 0.0000 / 0.9404 ± 0.0052 | 1.0000 ± 0.0000 / 0.9487 ± 0.0049 | 0.9426 ± 0.0020 / 0.9095 ± 0.0071 | 0.9964 ± 0.0006 / 0.9391 ± 0.0054 | 1.0000 ± 0.0000 / 0.9467 ± 0.0052 | 1.0000 ± 0.0000 / 0.9359 ± 0.0062 |
| **MCC (Train/Test)** | 0.9980 ± 0.0009 / 0.9251 ± 0.0118 | 1.0000 ± 0.0000 / 0.8128 ± 0.0169 | 1.0000 ± 0.0000 / 0.8364 ± 0.0161 | 0.8339 ± 0.0065 / 0.7398 ± 0.0198 | 0.9893 ± 0.0018 / 0.8061 ± 0.0181 | 1.0000 ± 0.0000 / 0.8308 ± 0.0168 | 1.0000 ± 0.0000 / 0.8091 ± 0.0185 |

#### Table S3. eQTLs identified in three driver genes by MatrixeQTL

| SNP* | rs number | gene | beta | t-stat | *P*-value | FDR |
|---|---|---|---|---|---|---|
| chr8:38996464-C-A | rs7840270 | *ADAM9* | -0.560481 | -4.461647 | 0.000034 | 0.038072 |
| chr8:38997543-G-T | rs7831735 | *ADAM9* | -0.565580 | -4.359599 | 0.000049 | 0.046521 |
| chr19:7742229-G-A | rs11465401 | *MCEMP1* | 1.912424 | 4.333792 | 0.000054 | 0.048775 |
| chr19:7742364-G-A | rs11465397 | *MCEMP1* | 1.912424 | 4.333792 | 0.000054 | 0.048775 |
| chr11:60510522-C-T | rs189755275 | *MS4A4A* | 2.328040 | 4.358676 | 0.000049 | 0.046648 |
| chr11:60547398-G-A | rs76847438 | *MS4A4A* | 2.328040 | 4.358676 | 0.000049 | 0.046648 |
| chr11:60582964-G-A | rs10736707 | *MS4A4A* | -2.328040 | -4.358676 | 0.000049 | 0.046648 |
| chr11:60623519-G-A | rs10792287 | *MS4A4A* | -2.328040 | -4.358676 | 0.000049 | 0.046648 |

*The positions refer to GRCh38

**Figure S1. Type I interferon response**

**A.** Interferon stimulated genes (ISG) scores based on the mean normalized expression of six genes (*IFI44L*, *IFI27*, *RSAD2*, *SIGLEC1*, *IFIT1*, and *ISG15*) obtained from RNAseq data. **B.** Heatmap showing the expression of type I IFN-related genes obtained from RNAseq data. Upregulated proteins are shown in red, and downregulated proteins are shown in light blue. **C.** IFNα2a (pg/ml) concentration evaluated using an ultra-sensitive S-PLEX Human IFNα2a Kit (Mesoscale Discovery). **D.** Time-dependent IFNα2a concentration in the critical group. **E.** Quantification of plasmacytoid dendritic cells as a percentage of PBMCs. The *P*-values were determined with the Kruskal-Wallis test followed by Dunn's posttest for multiple group comparisons; $*P < 0.05$, $** P < 0.01$, $*** P < 0.001$, $**** P < 0.0001$.

**Figure S2. Additional immune profiling of healthy individuals, noncritical and critical COVID-19 patients by mass cytometry**

Proportions of modified lymphocyte subsets from COVID-19 patients and healthy controls as determined by mass cytometry. The proportions of dendritic cell subsets (**A**), monocyte subsets (**B**), NK cell subsets (**C**), NKT cells (**D**), γδ T cells (**E**) and granulocyte subsets (traces), including neutrophils (**F**), are shown. Each dot represents a single patient. The *P*-values were determined with the Kruskal-Wallis test, followed by Dunn's posttest for multiple group comparisons; $* P < 0.05$, $** P < 0.01$, $*** P < 0.001$, $**** P < 0.0001$.

**Figure S3. SARS-CoV-2 immunization in healthy individuals, non-critical and critical COVID-19 patients**

**A.** Antibody reactivities against three viral antigens expressed in mean fluorescence intensities (MFI). *P*-values were determined with the Kruskal-Wallis test, followed by Dunn's post-test for multiple group comparison; $* P < 0.05$, $** P < 0.01$, $*** P < 0.001$, $**** P < 0.0001$. **B.** Half-maximal neutralizing titer (NT50) values of SARS-CoV-2 pseudo-viruses. *P*-values were determined with the Mann-Whitney's test; $* P < 0.05$, $** P < 0.01$, $*** P < 0.001$, $**** P < 0.0001$. **C.** Neutralization activity of type IFN autoantibodies was assessed by measuring the IFN stimulated response after treatment of HEK293T cells with patient or control serum. Relative luciferase activity is shown after stimulation with 10 ng/ml of IFNα or IFNω. ISRE, IFN stimulation response element; RLU, relative light units. The dotted line indicates the cutoff for positivity. For individuals in which different time points were measured, the lowest RLU value is shown. **D.** Time-course evaluation of the anti-SARS-CoV-2 neutralizing activity (left axis) and the anti IFN type I autoantibodies (right axis) in two critical patients with anti-type I autoantibodies. The farmost right points correspond to the time points represented in panel C.

**Figure S4. Results of *in silico* perturbation experiments**

Left: change in BIC (Bayesian Information Criterion) when perturbing each gene individually. Genes are ordered by the number of ancestors minus the number of descendants for the DAG shown in Figure 5B: the top five driver genes are the five leftmost points, and the top five response genes are the five rightmost points. Right: Change in the BIC of a random sample of five genes from the left. The mean BIC of the top five driver genes is shown in red.

**Figure S5. Validation of the RNA-seq signature-based classification performance of critical and recovered critical COVID-19 patients**

**A.** ROCs on the train and test sets for critical vs recovered critical groups comparison in the replication cohort using the 600 gene signature identified from the initial cohort. All methods performed similarly. **B.** Classification metrics. **C.** Box plots showing the normalized gene counts of the five driver genes in critical and recovered critical patients. The indicated values correspond to the FDR.

**Figure S6. *ADAM9* expression in publicly available data**

Box plots showing the normalized gene counts of *ADAM9* in healthy (n=17), severe (n=8) and ICU (n=3) patients in the GSE152418 dataset reported by Arunachalam et al., in Science (DOI:10.1126/science.abc6261). The indicated values correspond to the FDR.

**Figure S7. Validation of *ADAM9* silencing**

**A.** Quantitative RT-PCR of the *ADAM9* transcript in Vero 76 or A549-ACE2 cells silenced with a control siRNA or an *ADAM9*-specific siRNA. The average silencing achieved was 66% and 93% for Vero 76 and A549-ACE2 cells, respectively (mean of three representative experiments). **B.** Western blot of Vero 76 and A549-ACE2 cells that were not transfected (NT), were silenced with a control siRNA (ctl) or were silenced with an ADAM9-specific siRNA (sil.).

### Figure S1.

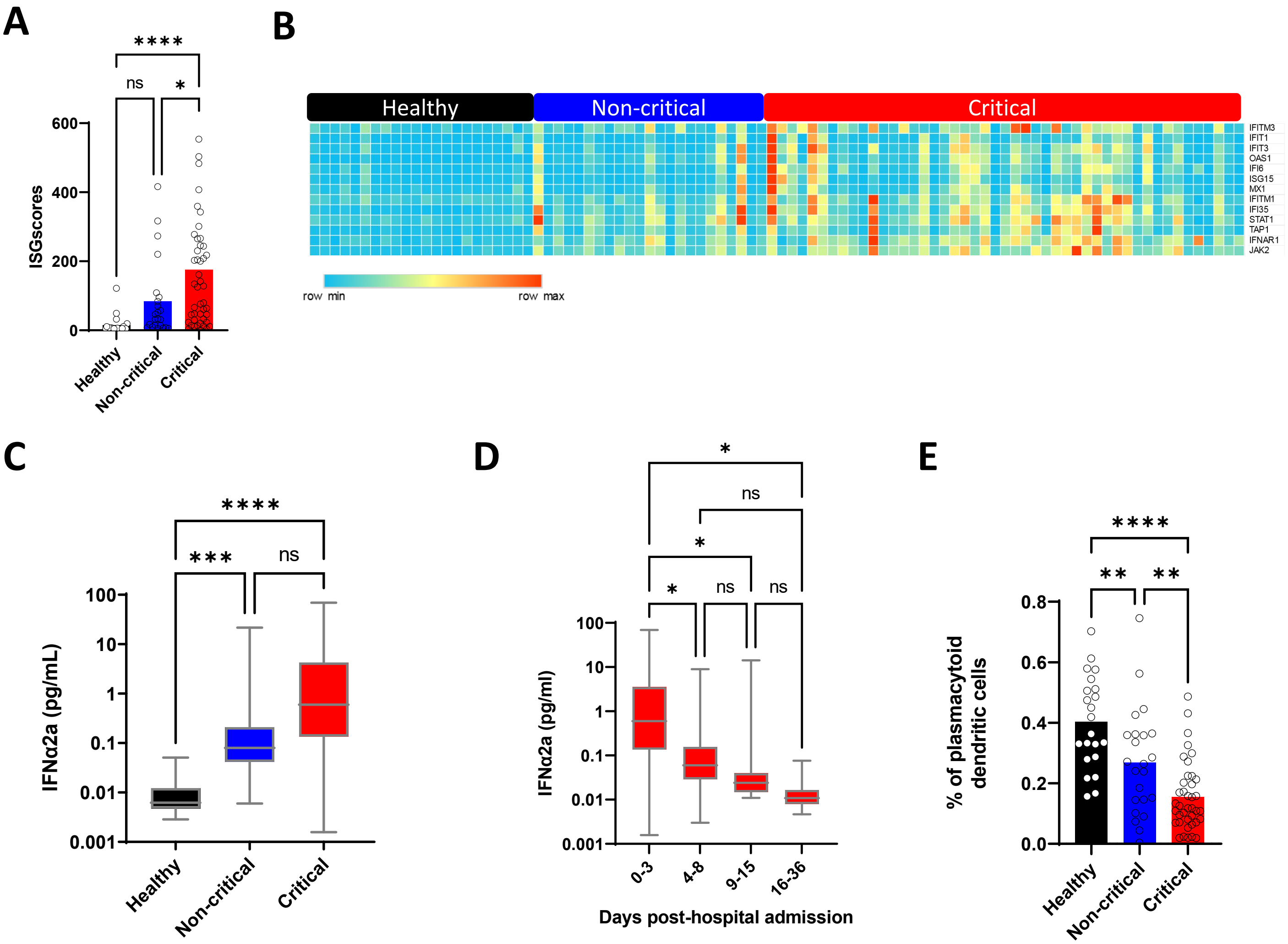
A
B
C
D
E
ISGscores
600
400
200
0
ns
*
****
Healthy
Non-critical
Critical
IFITM3
IFIT1
IFIT3
OAS1
IFI6
ISG15
MX1
IFITM1
IFI35
STAT1
TAP1
IFNAR1
JAK2
row min
row max
IFNα2a (pg/mL)
100
10
1
0.1
0.01
0.001
***
IFNα2a (pg/ml)
0-3
4-8
9-15
16-36
Days post-hospital admission
% of plasmacytoid dendritic cells
0.8
0.6
0.4
0.2
0.0
**

### Figure S2.

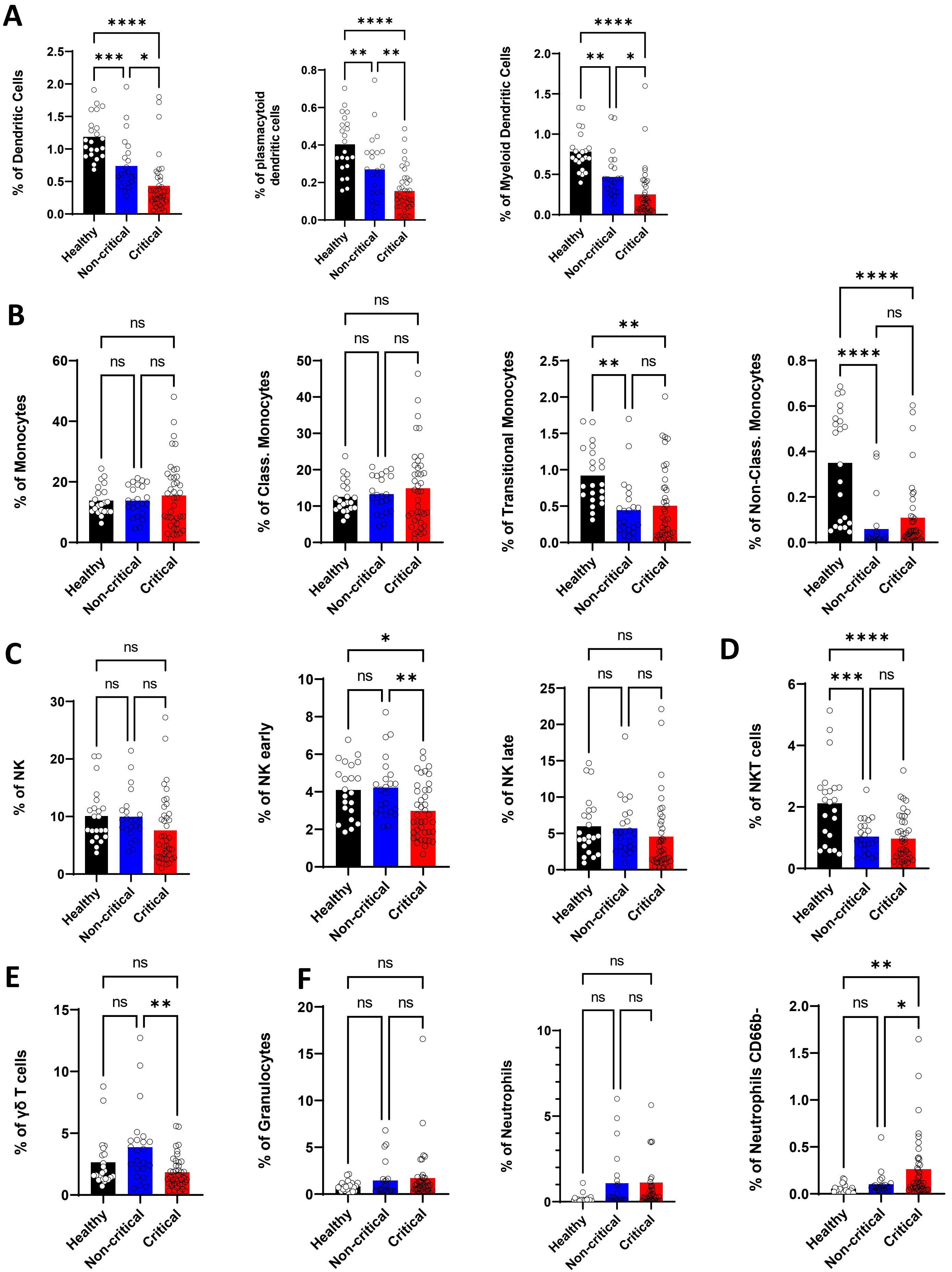
A
% of Dendritic Cells
% of plasmacytoid dendritic cells
% of Myeloid Dendritic Cells
B
% of Monocytes
% of Class. Monocytes
% of Transitional Monocytes
% of Non-Class. Monocytes
C
% of NK
% of NK early
% of NK late
D
% of NKT cells
E
% of γδ T cells
F
% of Granulocytes
% of Neutrophils
% of Neutrophils CD66b-
Healthy
Non-critical
Critical
ns
*
**
***
****

### Figure S3.

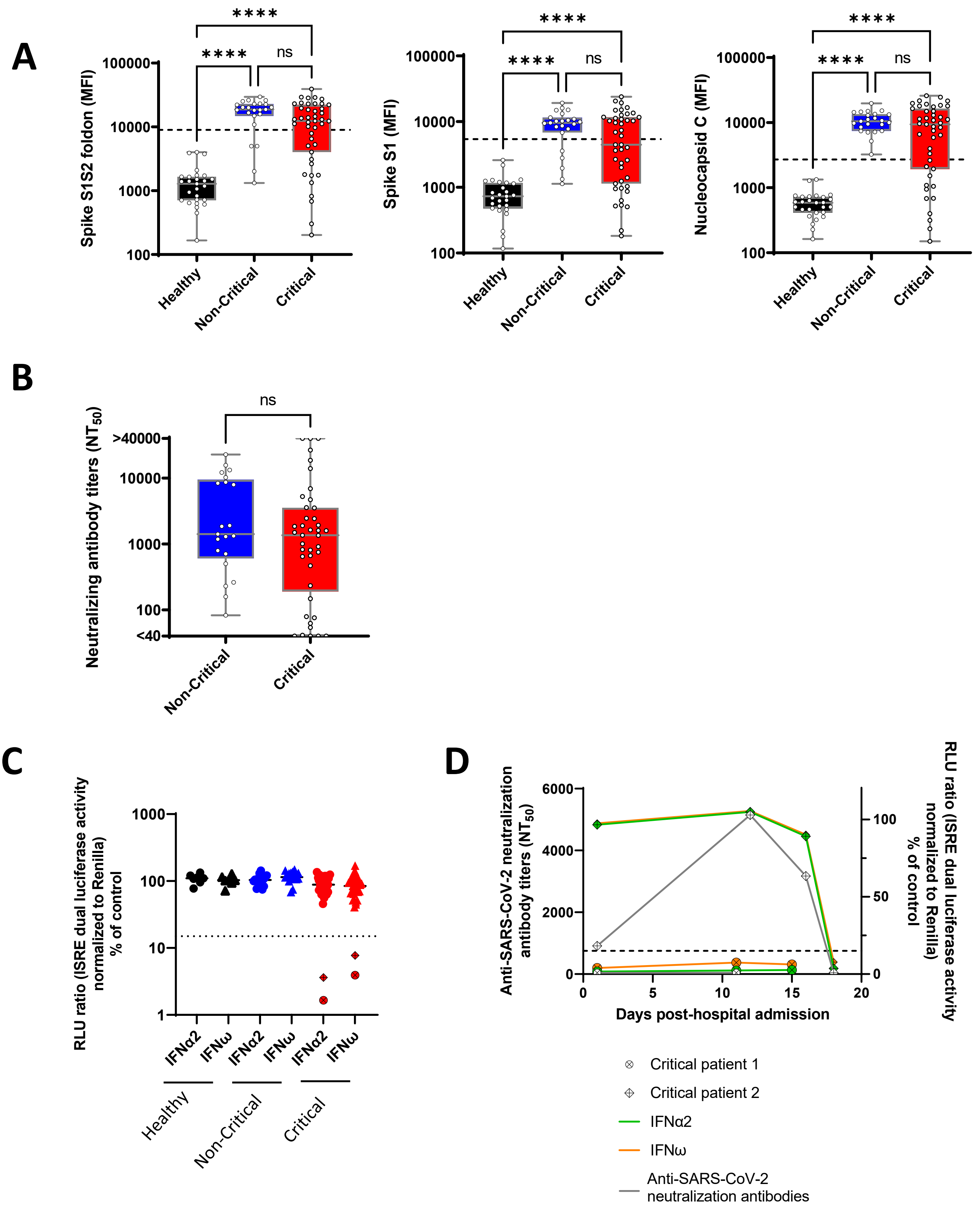

A
Spike S1S2 foldon (MFI)
Spike S1 (MFI)
Nucleocapsid C (MFI)
100000
10000
1000
100
****
ns
Healthy
Non-Critical
Critical
B
Neutralizing antibody titers ($NT_{50}$)
>40000
10000
1000
100
<40
ns
C
RLU ratio (ISRE dual luciferase activity normalized to Renilla) % of control
IFNα2
IFNω
Healthy
Non-Critical
Critical
D
Anti-SARS-CoV-2 neutralization antibody titers ($NT_{50}$)
RLU ratio (ISRE dual luciferase activity normalized to Renilla) % of control
Days post-hospital admission
Critical patient 1
Critical patient 2
IFNα2
IFNω
Anti-SARS-CoV-2 neutralization antibodies

### Figure S4.

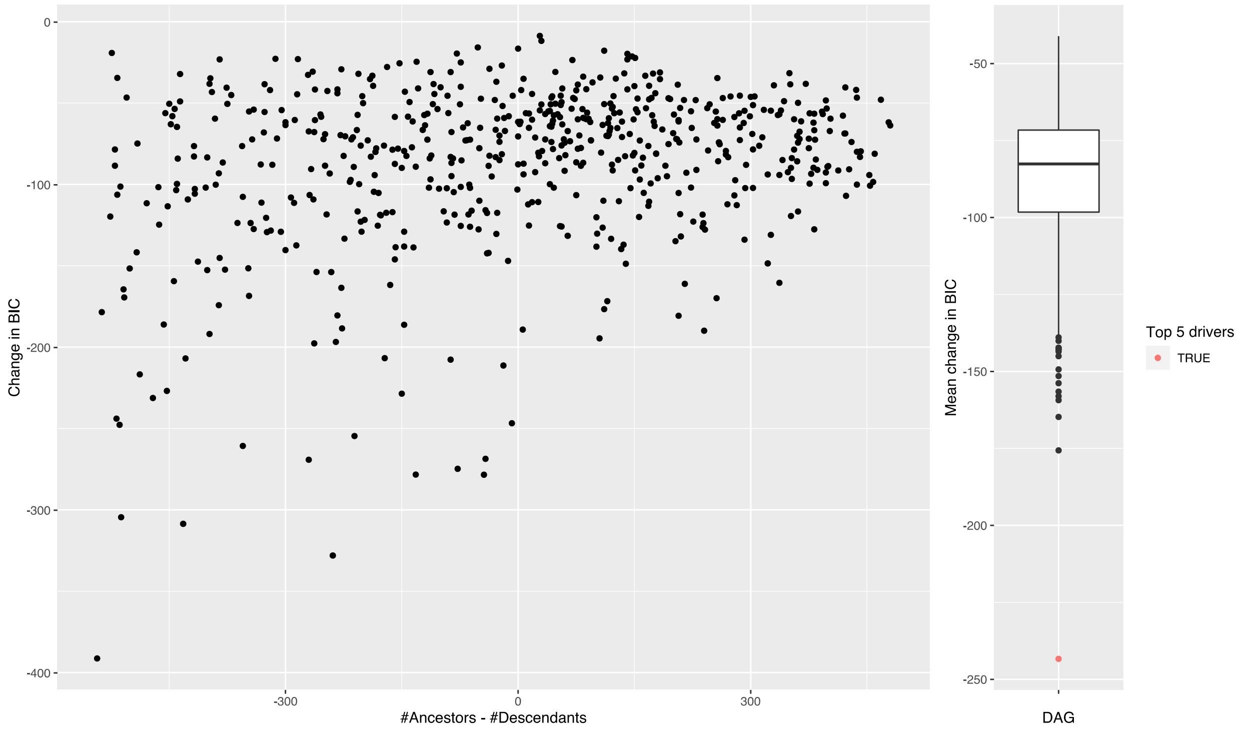
Change in BIC
0
-100
-200
-300
-400
-300
0
300
#Ancestors - #Descendants
Mean change in BIC
-50
-100
-150
-200
-250
DAG
Top 5 drivers
TRUE

### Figure S5.

A

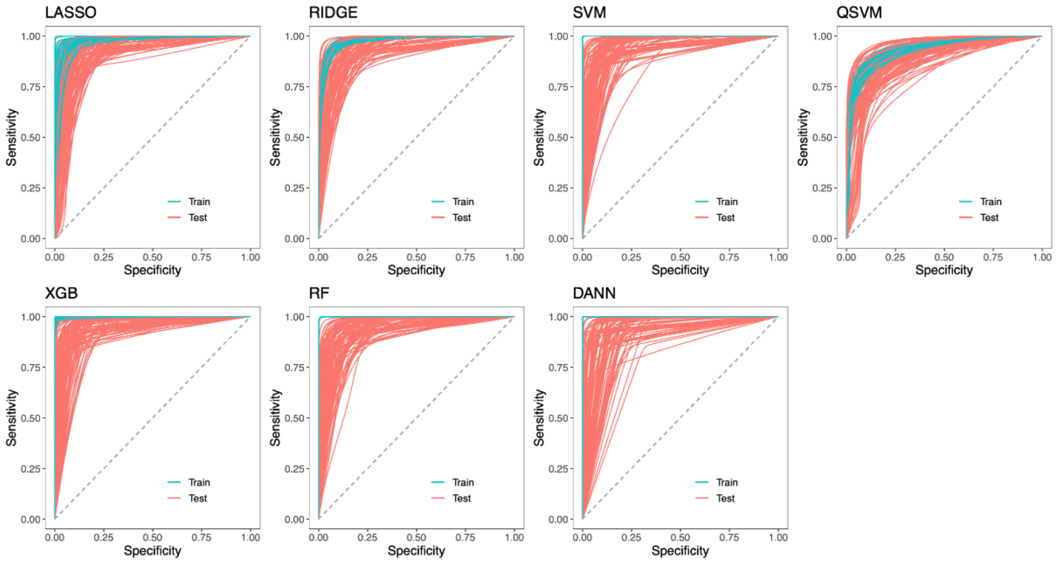


B

| | LASSO | Ridge | SVM | qSVM | XGB | RF | DANN |
|---|---|---|---|---|---|---|---|
| Accuracy (Train/Test) | 0.9535 ± 0.0016 / 0.9233 ± 0.0044 | 0.9453 ± 0.0010 / 0.9123 ± 0.0055 | 1.0000 ± 0.0000 / 0.9160 ± 0.0047 | 0.9077 ± 0.0019 / 0.8873 ± 0.0064 | 0.9983 ± 0.0004 / 0.9223 ± 0.0045 | 1.0000 ± 0.0000 / 0.9180 ± 0.0049 | 1.0000 ± 0.0000 / 0.9130 ± 0.0051 |
| Balanced Acc. (Train/Test) | 0.9544 ± 0.0016 / 0.9262 ± 0.0042 | 0.9463 ± 0.0010 / 0.9154 ± 0.0053 | 1.0000 ± 0.0000 / 0.9184 ± 0.0045 | 0.9090 ± 0.0018 / 0.8894 ± 0.0062 | 0.9983 ± 0.0004 / 0.9242 ± 0.0044 | 1.0000 ± 0.0000 / 0.9197 ± 0.0048 | 1.0000 ± 0.0000 / 0.9156 ± 0.0049 |
| AUROC (Train/Test) | 0.9849 ± 0.0009 / 0.9628 ± 0.0031 | 0.9879 ± 0.0003 / 0.9684 ± 0.0029 | 1.0000 ± 0.0000 / 0.9671 ± 0.0029 | 0.9548 ± 0.0011 / 0.9486 ± 0.0041 | 0.9999 ± 0.0000 / 0.9789 ± 0.0023 | 1.0000 ± 0.0000 / 0.9644 ± 0.0032 | 1.0000 ± 0.0000 / 0.9643 ± 0.0033 |
| F1 (Train/Test) | 0.9547 ± 0.0016 / 0.9233 ± 0.0046 | 0.9466 ± 0.0010 / 0.9115 ± 0.0058 | 1.0000 ± 0.0000 / 0.9164 ± 0.0050 | 0.9061 ± 0.0019 / 0.8849 ± 0.0063 | 0.9984 ± 0.0004 / 0.9235 ± 0.0046 | 1.0000 ± 0.0000 / 0.9199 ± 0.0049 | 1.0000 ± 0.0000 / 0.9133 ± 0.0053 |
| MCC (Train/Test) | 0.9080 ± 0.0032 / 0.8547 ± 0.0081 | 0.8917 ± 0.0021 / 0.8346 ± 0.0102 | 1.0000 ± 0.0000 / 0.8406 ± 0.0087 | 0.8179 ± 0.0036 / 0.7839 ± 0.0121 | 0.9966 ± 0.0008 / 0.8515 ± 0.0085 | 1.0000 ± 0.0000 / 0.8418 ± 0.0094 | 1.0000 ± 0.0000 / 0.8343 ± 0.0096 |

C

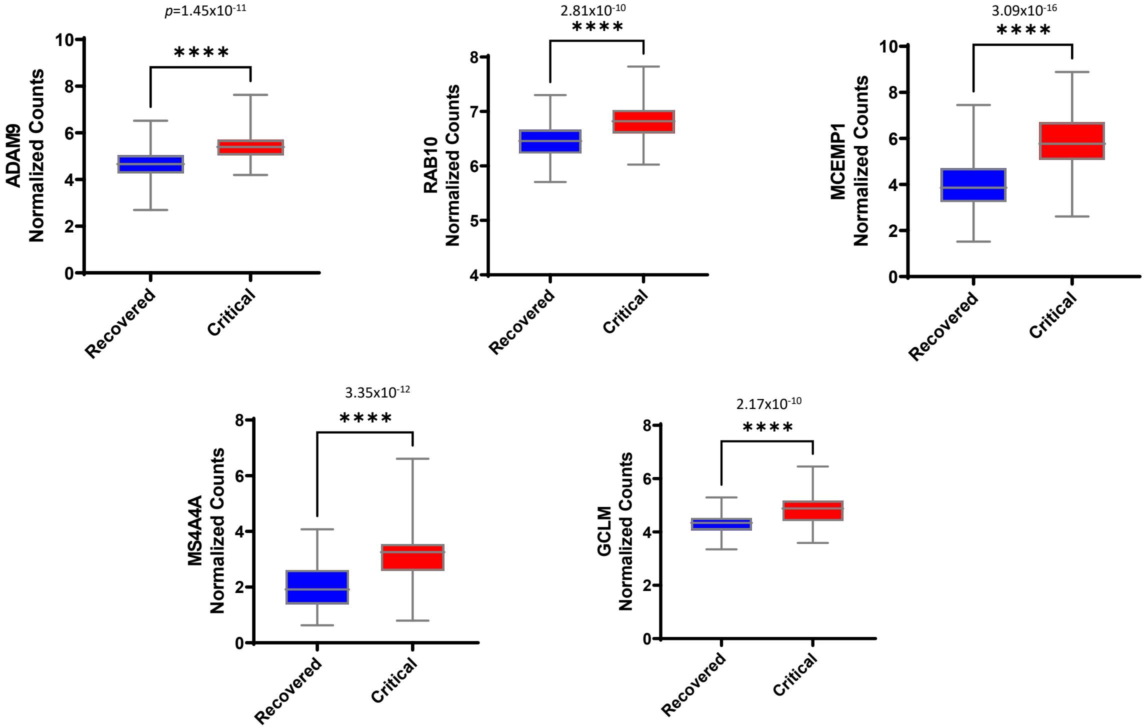

**Figure S6.**

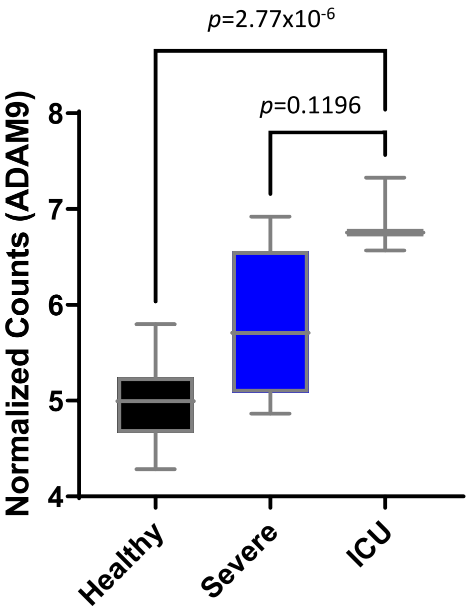

p=2.77x10-6
p=0.1196
Normalized Counts (ADAM9)
8
7
6
5
4
Healthy
Severe
ICU

**Figure S7.**

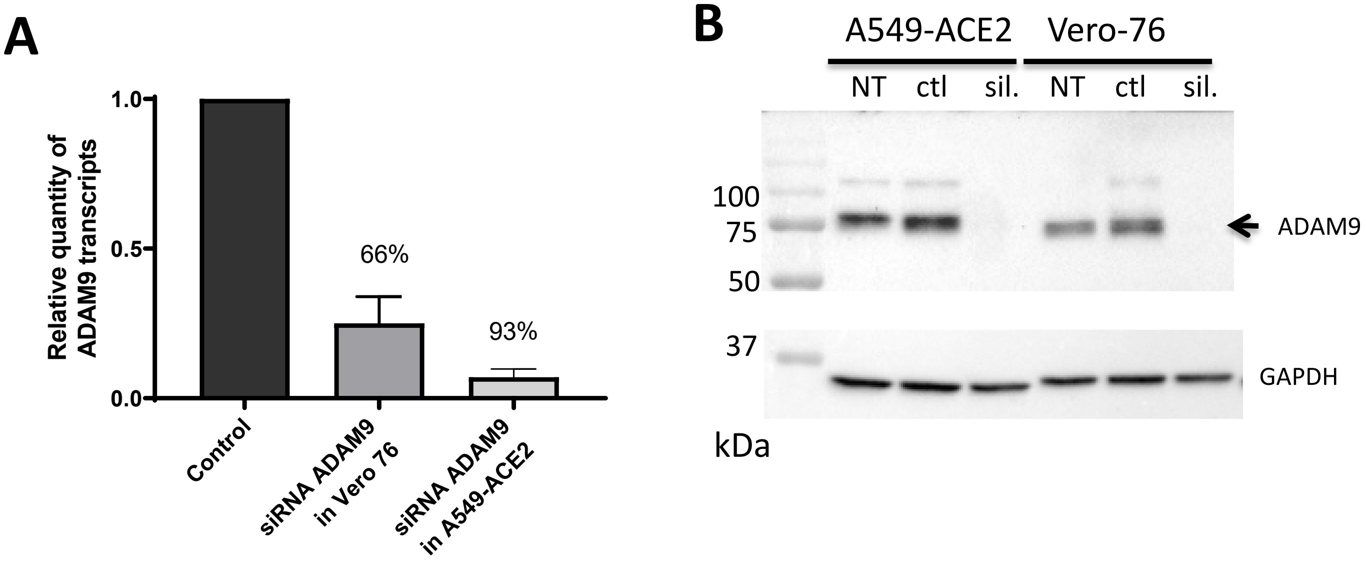
A
Relative quantity of ADAM9 transcripts
1.0
0.5
0.0
66%
93%
Control
siRNA ADAM9 in Vero 76
siRNA ADAM9 in A549-ACE2
B
A549-ACE2
Vero-76
NT
ctl
sil.
NT
ctl
sil.
100
75
50
37
ADAM9
GAPDH
kDa
